# Prolonged grief during and beyond the pandemic: Factors associated with levels of grief in a four time-point longitudinal survey of people bereaved in the first year of the COVID-19 pandemic

**DOI:** 10.1101/2023.06.22.23291742

**Authors:** Emily Harrop, Renata Medeiros Mirra, Silvia Goss, Mirella Longo, Anthony Byrne, Damian JJ Farnell, Kathy Seddon, Alison Penny, Linda Machin, Stephanie Sivell, Lucy E Selman

## Abstract

**Background:** The COVID-19 pandemic has been a devastating and enduring mass-bereavement event, with uniquely difficult sets of circumstances experienced by people bereaved at this time. However, little is known about the long-term consequences of these experiences, including the prevalence of Prolonged Grief Disorder (PGD) and other conditions in pandemic-bereaved populations.

**Methods:** A longitudinal survey of people bereaved in the UK between 16 March 2020 and 2 January 2021, with data collected at baseline (n=711), c. 8 (n=383), 13 (n=295) and 25 (n=185) months post-bereavement. Using measures of Prolonged Grief Disorder (PGD) (Traumatic Grief Inventory), grief vulnerability (Adult Attitude to Grief Scale), and social support (Inventory of Social Support), this analysis examines how participant characteristics, characteristics of the deceased and pandemic-related circumstances (e.g. restricted visiting, social isolation, social support) are associated with grief outcomes, with a focus on levels of PGD.

**Results:** At baseline, 628 (88.6%) of participants were female, with a mean age of 49.5 (SD 12.9). 311 (43.8%) deaths were from confirmed/suspected COVID-19. Sample demographics were relatively stable across time points 34.6% of participants met the cut-off for indicated PGD at c. 13 months bereaved and 28.6% at final follow-up. Social isolation and loneliness in early bereavement and lack of social support over time strongly contributed to higher levels of PGD, whilst feeling well supported by healthcare professionals following the death was associated with reduced levels of PGD. Characteristics of the deceased most strongly associated with lower PGD scores, were a more distant relationship (e.g. death of a grandparent), an expected death and death occurring in a care-home. Participant characteristics associated with higher levels of PGD included low level of formal education and existence of medical conditions.

**Conclusion:** Results suggest higher than expected levels of PGD compared with pre-pandemic times, with important implications for bereavement policy, provision and practice now (e.g. strengthening of social and specialist support) and in preparedness for future pandemics and mass-bereavement events (e.g. guidance on infection control measures and rapid support responses).

## BACKGROUND

Millions of people were bereaved during the COVID-19 pandemic, with close to seven million reported deaths caused by the virus world-wide, and over 200,000 in the UK (1). This prolonged mass-bereavement event was characterised by high death-rates and unprecedented restrictions to usual end-of-life, death and mourning practices and social life in general. In the early months of the pandemic, observers predicted worsened grief and bereavement outcomes in response to the sudden and unexpected nature of COVID-19 deaths, the traumatic circumstances in which these deaths occurred and the likely diminished coping capacities of bereaved people (and the people and services supporting them) (2–4).

More than three years on from the start of the global pandemic, there is now a considerable body of evidence documenting the impacts of these devastating, unique sets of circumstances on those bereaved at this time. However, little is currently known about the longer-term consequences of pandemic bereavement, including which groups of people are most at risk of adverse outcomes over time and whether initial predictions of increased levels of Prolonged Grief Disorder (PGD) are substantiated (2–4). Essential characteristics of PGD include persistent and pervasive longing for, or preoccupation with, the deceased, associated with intense emotional pain (e.g., sadness, guilt, denial), functional impairment, and atypically prolonged symptoms relative to cultural norms (and a minimum of six months post-bereavement) (5,6). Although figures on PGD or complex grief vary between studies (e.g. between 6 and 20%; 7-10) in non-pandemic public health models, it is commonly accepted that around 10% of bereaved people will experience PGD, requiring specialist psychological intervention, while those with ‘moderate’ level needs and risk (estimated at around 30%) may also need formal bereavement support such as peer-support groups or grief counselling (7,11). Evidence on PGD levels during and following the pandemic is therefore needed to better understand the long-term grief and associated support-needs of people bereaved during this and future pandemics, with implications for bereavement service-planning and delivery. It is also critical for informing policy considerations relating to infection-control isolation measures in both the current COVID-19 recovery phase, and as part of our preparedness for future outbreaks of infectious diseases.

Most evidence to date on the grief and mental health consequences of pandemic bereavement is from studies conducted in China (12,13), North America (14–18), Holland (4,19,20) and the earlier qualitative and quantitative results from this UK-based study (21–26). Several of these cross-sectional studies (including our baseline publication, 23) indicate higher levels of grief and functional impairment amongst people bereaved during the pandemic, compared with pre-pandemic populations (14–20). Many of these studies have demonstrated the negative grief impacts of pandemic-specific or related circumstances. These have included restricted visiting at the end of life and opportunities to say goodbye (15,17,20,24), sub-optimal communication and support from healthcare staff at the end of life (17,22,24), disrupted funerals (17, 23,24), experiences of loneliness and isolation (6,15,17,23,24) and the role of disrupted meaning-making in mediating the effects of these sets of circumstances (15,17). Another study, by contrast, found no differences in levels of PGD, attendance at, or evaluations of funerals and other mourning rituals, between pandemic and pre-pandemic bereaved populations (28).

Of particular interest early on in the pandemic was whether COVID-19 deaths would be associated with worse grief experiences than other types of death. Higher levels of grief and other psychological conditions have been identified amongst those bereaved by COVID-19 than would be expected in non-pandemic populations (12-14,17), or compared with ‘natural’ but not ‘unnatural’ causes of deaths pre and during the pandemic (4,18–20), with the ‘unexpected’ nature of these deaths an explanatory factor (19,20). However, other studies (including the baseline results from this study) have not found significant differences in grief and other psychological outcomes between COVID-19 and non-COVID-19 bereavement during the pandemic (15,23). Several of these studies have also investigated the effects of demographic and other known risk factors for adverse grief and health outcomes. Consistent with pandemic (12-14,27,29) and non-pandemic research (7,30), our baseline results identified relationship with the deceased as the strongest factor predicting grief vulnerability (23). Younger age of the deceased was also associated with worse baseline grief vulnerability (23), as in other studies (7,29,31). Age, gender, race/ethnicity of the bereaved person, and time since death were not significantly associated with level of grief in our baseline results (23) or functional impairment in one of the US studies (14). By contrast, lower levels of education were associated with poorer outcomes in our baseline results (23), reflecting the findings of previous non-pandemic research (32–34). Related associations with low income have also been identified, including a study involving pandemic and pre-pandemic bereaved participants (7).

However, to date no longitudinal results have been published on grief outcomes during the pandemic, and most of the above mentioned studies included pandemic-bereaved populations who were on average bereaved less than six months before (e.g. 4,12,14,15,17,19,20,23), thus limiting observations that can be made regarding levels of, and factors associated with, PGD levels in their respective populations. Addressing this knowledge gap, this paper reports longitudinal results regarding factors associated with PGD scores amongst a cohort of participants bereaved during the first two waves of the pandemic in the UK, using data collected at four time-point survey rounds, up to 25 months post-bereavement.

## METHODS

### Study design and aim

A longitudinal survey of people bereaved during the pandemic in the UK. The web-based survey was conducted as part of a larger mixed methods study, which aimed to investigate the grief experiences, support needs and use of bereavement support by people bereaved during the pandemic (21–26). The current analysis examines how clinical and demographic factors, and pandemic-related challenges are associated with levels of indicated PGD in a cohort of participants surveyed at baseline (T1) and c. 8, 13 and 25 months post-bereavement (T2-T4). The mediating role of perceived social support was also investigated in this analysis, reflecting its established importance for healthy grieving and adaptation (e.g. 10, 35-37) and its likely association with other demographic factors potentially also predictive of grief severity (e.g. gender, ethnicity, age).

The Checklist for Reporting Results of Internet E-Surveys (38) was followed.

### Survey development

An open web survey was designed by the research team, which includes a public representative (KS), with input from the study advisory group. Each survey was piloted, refined with public representatives with experience of bereavement, and tested by the study advisory group and colleagues. Non-randomised open and closed questions covered end of life experiences, grief experiences, and perceived needs for, access to and experiences of formal and informal bereavement support (21,22).

### Outcome measures

Prolonged Grief Disorder was assessed at surveys T2-T4 using the Traumatic Grief Inventory Self-Report version (TGI-SR) (39,40). This widely used 18-item self-report measure assesses symptoms of Persistent Complex Bereavement Disorder (PCBD) and PGD, as defined by Prigerson et al. (9). The TGI-SR includes all 16 symptoms of PCBD, one additional symptom of PGD that is not part of the PCBD criteria (i.e. item 12: “feeling stunned/shocked”) and one item tapping “functional impairment” (i.e. item 13), included in criteria-sets for both PCBD and PGD (40). Participants rated the frequency of symptoms (e.g., “I felt a strong longing or yearning for the deceased”) during the previous month on 5-point scales (1 = never and 5 = always). Total scores ranged from 16–80. A cut- off score of ≥54 (i.e., mean item score of 3.0) is indicative of PCBD and PGD when using the total score (40). The measure was not used at baseline as PGD should be assessed at least six months after a death, and PCBD at least 12 months afterwards (39,40).

*Vulnerability in Grief* was assessed in all survey time points using the validated 9-item Adult Attitude to Grief (AAG) scale (41), with our reasons for selecting this measure reported in baseline publications (21,23). The scale is based on the Range of Response to Loss model (42), which identifies three distinct responses: being ‘overwhelmed’, a state dominated by emotional/cognitive distress; being ‘controlled’, needing to avoid emotional expression and focus on day-to-day life; and being balanced or ‘resilient’, feeling supported and able to cope. AAG subscale scores indicate levels of feeling overwhelmed, controlled, and reversed resilience on a scale of 0 (none) to 12 (very high). An overall index of vulnerability (IOV) is calculated by summing subscale scores (IOV: 0–20 = low vulnerability, 21–23 = high vulnerability, and 24–36 = severe vulnerability (41). Although this analysis and publication is focused on PGD, we included this measure to enable comparisons to be made with our analysis of baseline survey data, which did not include the TGI measure (23).

*Social support* was assessed using the Inventory of Social Support (ISS) (43). The ISS is a 5-item measure that assesses how far a bereaved person can talk with other people about their loss in a way which supports adaptive coping. The measure includes such statements as “I can express my feelings about my grief openly and honestly” “There is at least one person I can talk to about my grief.” Participants respond to such statements on a 5-point Likert scale that ranges from 1 (Does not describe me very well) to 5 (Describes me very well). Higher scores on the ISS indicate higher levels of social support. Social support was investigated both as a dependent variable and as an independent variable in the PGD/TGI model.

### Associated factors

We assessed whether participant characteristics and characteristics of the deceased, experiences of end-of-life care and pandemic-related problems independently predicted levels of PGD and IoV and whether perceived social support mediated the relationships of these variables and PGD scores. Factors included in the analysis are recognised risk factors for poor bereavement outcomes (age of deceased and bereaved, gender, time since death, relationship to deceased, expectedness of the death, ability to say goodbye to the deceased, support from healthcare professionals at the end of life, perceived social support) (10, 23,44–46) or are known to be indirectly associated with such outcomes (qualifications, health status, place of death, cause of death) (47,48).

*Pandemic-related problems:* Six items at baseline assessed pandemic-related challenges prior to and after the death, e.g. being unable to visit the person who died prior to their death, restricted funeral arrangements, social isolation and loneliness. All items were answered yes/no. Respondents were asked to tick all experiences that applied to them.

See supplementary file 1 for baseline questionnaire and supplementary file 2 for final questionnaire, including all measures used in this analysis.

### Study procedure

The baseline survey was administered via JISC (https://www.onlinesurveys.ac.uk/) and was open from 28th August 2020 to 5th January 2021 (21–24). It was disseminated to a convenience sample from social and mainstream media and via voluntary sector associations and bereavement support organisations, including those working with ethnic minority communities. Organisations helped disseminate the voluntary (non-incentivised) survey by sharing on social media, web-pages, newsletters, on-line forums and via direct invitations to potential participants. For ease of access, the survey was posted onto a bespoke study-specific website with a memorable URL (www.covidbereavement.com). Hard-copy postal surveys were available on request. The second, third and fourth follow up surveys were sent to baseline participants who consented to receive follow up surveys around seven, 13 and 25 months post date of death. These were personalised for each participant using individual survey links, labelled with their participant study IDs. Where baseline surveys were completed at least five months post-death (or the date of death was not given), the second survey was sent out two months after the first survey was received. All second-round surveys were completed between 20/11/20 and 24/08/2021 and on average 242 days (median = 234 days or 8 months) after the date of death (range 145 to 345 days). All third-round surveys were completed between 04/05/2021 and 09/01/22 and on average 408 days (median = 404 or 13 months) after the date of death (range 396 to 481 days). All fourth round surveys were completed between 17/05/2022 and 12/01/2023, on average 776 days (median = 774 or 25 months) after the date of death (range 762 days to 812 days).

Inclusion criteria for study enrolment: aged 18+; family member or close friend bereaved since social-distancing requirements were introduced in the UK (16/03/2020); death occurred in the UK; ability to consent. The initial section of the survey requested informed consent and details data protection.

### Data analysis

All analysis was performed using R (version 4.1.1, R Core Team, 2021), implemented in R-Studio (www.r-studio.com) (49). Descriptive statistics were used to describe all variables. The main outcome variable of interest in this study was levels of PGD, assessed through the TGI questionnaire. ISS scores were used both as a predictor of levels of PGD and as an outcome variable, indirectly exploring the potential mediation effect of ISS between some of the independent variables and PGD levels. Since PGD and ISS scores were not collected at baseline, IOV scores for the AAG questionnaire were also used as outcome variable to allow for comparisons between baseline and the other time points. Missing values in the AAG questionnaire were inputted for each sub-category if two of the three scores were available, by taking the mean of the two; IOV scores were used only if data were available for all three sub-categories. Missing data for ISS and TGI scores were inputted by using the mean of the remaining scores; the maximum number of TGI or ISS imputations for the same participant was three. The thresholds used for IOV categories followed Sim et al (41) and the threshold used for PCBD and PGD was TGI score ≥54 (40).

Independent variables were classified into three categories: characteristics of the participant/bereaved, characteristics of the deceased, and characteristics of the experience of bereavement. The latter included the six items assessing the pandemic-related challenges prior to and after the death as well as whether participants felt well supported by healthcare professionals immediately after the death. For the analysis of PGD levels, ISS was also included in this category.

Independent variables with more than 5% missing data at any of the four time points were not considered for analysis. A summary analysis of missing data was carried out for the remaining variables to check if there were any obvious patterns of missingness. In order to maximise the sample, results presented are from analysis carried on all data available for each variable or combination of variables used in each analysis, but all the analyses were also carried out using complete cases (i.e. excluding any participants with at least one missing data for any of the variables of interest) as a control to ensure that the missing data were not causing a great influence in the results.

Days since bereavement was scaled into z-scores due to the wide range of values. This variable was initially tested both as a linear and as a quadratic term, but the latter showed very small effects and did not improve the fit of the statistical models significantly and hence the scaled linear effect was used instead. Other genders besides male or female were not included in the analysis due to the very small sample sizes. For relationship with the deceased, the categories ‘other family member’ and ‘colleague or friend’ were merged into one category in the analysis. Likewise, for place of death, the categories “other” and “don’t know” were also merged. The existence of any medical conditions, whether the bereaved respondent was unemployed during the pandemic and whether they had suffered any further bereavements throughout the study were considered cumulatively – i.e. if a participant had reported a medical condition, becoming unemployed or suffering a further bereavement in one round of the survey, that was carried through even though they might not have reported it again in a subsequent round.

The first step of the analysis consisted of fitting General Linear Mixed Models (GLMMs) to assess the single effects of each independent variable on each of the outcome variables (IOV, ISS and PGD levels). The second step consisted of fitting GLMMs for each group of variables in combination to assess which group of variables (characteristics of the participant/bereaved, characteristics of the deceased, or characteristics of the experience of bereavement) were better at explaining each of the outcome variables.

The third step consisted of fitting GLMMs that included all groups of variables in combination to assess the effect of the experience of bereavement in each of the outcome variables, while controlling for the characteristics of the participant/bereaved and the characteristics of the deceased. These models failed to converge and, hence, a Principal Component Analysis (PCA) was carried out on the six items assessing the pandemic-related challenges prior to and after the death, to assess if these could be reduced to a smaller number of factors. The models were initially run with the factors from the PCA, instead of the six different items. Variables with negligible effect sizes were then removed and the factors were replaced by the six items in the reduced model. The final models were used to compute and plot predictions showing the change of IOV, ISS and PGD levels across time for the different levels of the independent variables that showed medium or strong effect sizes. For PGD, two different models were fit, one with ISS as predictor and one without, to assess the role of ISS as a potential mediator of other predictors.

All models included participant ID as random term and days since bereavement as a covariate. Interactions between days since bereavement and all other variables were tested at this stage. Statistically significant interactions (p<0.05) found in the single models were also tested in the final models. Model estimates and standardise effect sizes were used to evaluate the effect of each variable independently on IOV, ISS and PDG levels; Cohen’s *d* were used for categorical predictors: *d* = 0.3: small effect, *d* = 0.5: medium effect, *d* = 0.8: large effect, *d* = 1.2: very large; and partial R for continuous predictors: <.10: trivial effect, 0.1 - 0.3: small to medium effect, 0.3 - 0.5: medium to large effect, >0.50: large to very large effect; 50). Where categorical predictors contained more than one group, we chose a reference category that allowed us to show the maximum difference in means between any two groups and the average standard deviation across all groups (i.e. maximum effect size of the difference). By using a standardised measure of effect size, the effects of factors on outcomes could be compared directly and patterns across multiple outcomes ascertained. R were used to assess the overall fit of the models in terms of their explanatory power and to explore which variables were the greatest contributors to explaining variability in IOV, ISS and PGD scores.

The fit of the models was assessed visually. Residuals were checked for normality and homoscedasticity and all the models showed a good enough fit. Correlation matrices showed no problems with multicollinearity.

The full list of R packages and functions used in the analysis is presented in supplementary file 3.

### Ethical approval

The study was approved by Cardiff University School of Medicine Research Ethics Committee (SMREC 20/59) and conducted in accordance with the Declaration of Helsinki. All respondents provided informed consent.

## RESULTS

A total of 711 participants answered the survey at baseline (T1), 383 answered it in the second round (T2) and 295 answered it in the third round (T3), including 35 who had not completed T2. 185 answered it on the fourth round (T4), two of whom had only completed T1 (but not T2 or T3) and 19 who had completed either T2 or T3. A total of 165 participants completed the survey at all time points.

### Characteristics of the participants

Table 1 shows the characteristics of the participants for each round of surveys. The average age of participants was around 50 and most participants were women, heterosexual and white. Across the four rounds of the study there was a tendency of the youngest and oldest participants to stop engaging, as well as those from minoritized ethnic backgrounds and those with lowest qualification levels. Over three quarters of the participants had not suffered unemployment or further bereavements at baseline, but towards the end of the study approximately 40% had experienced either circumstance. Similarly, approximately 40% of participants reported having medical conditions at baseline, but towards the end of the study, this increased to approximately 60% of participants. Overall, there was no strong change in participants’ demographic characteristics throughout the study. Spiritual/religious beliefs, sexual orientation and region were not considered for analyses due to missing data.

**Table 1.**
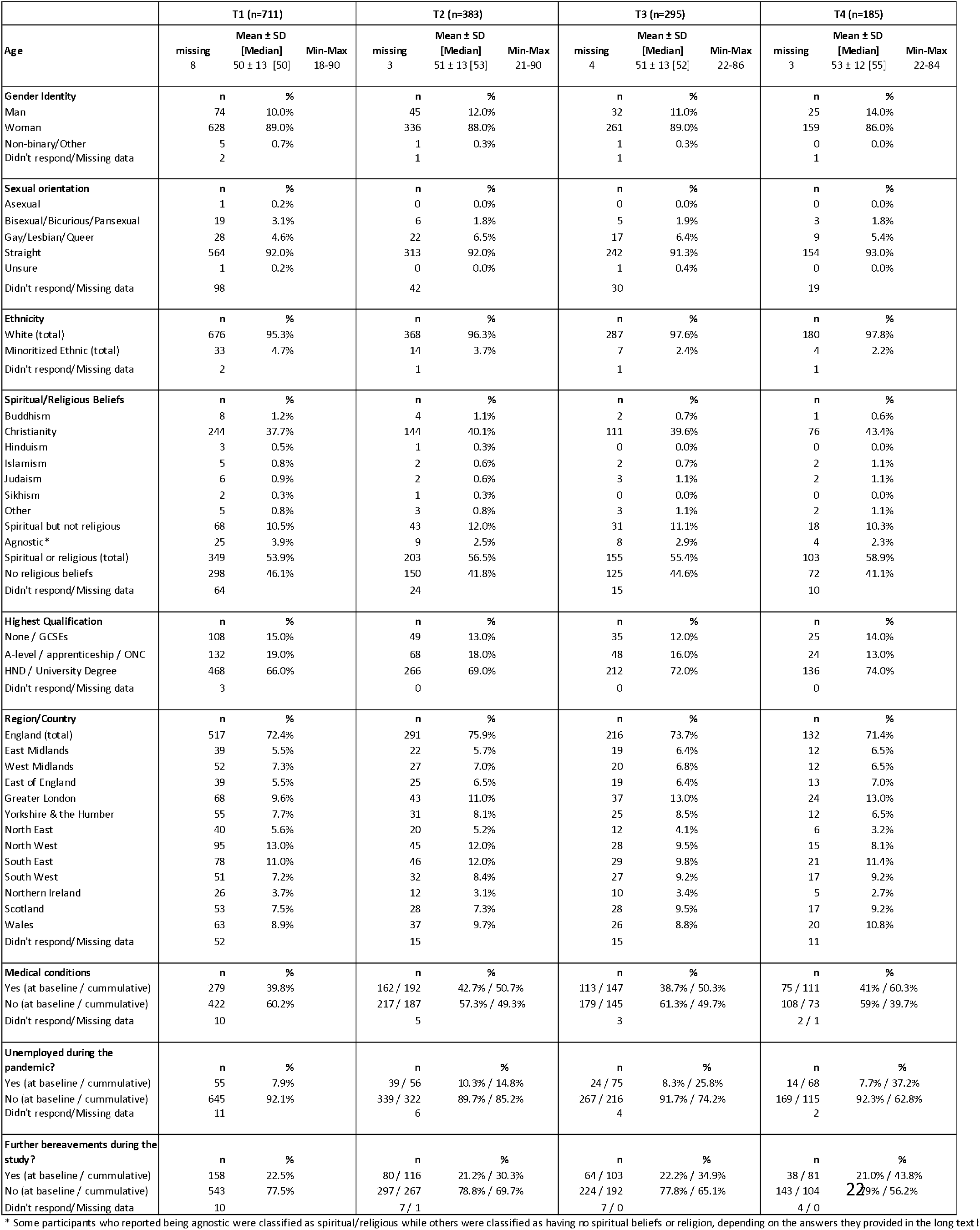
Characteristics of the study participants. All variables were taken at baseline only, except for medical conditions, unemployed since the pandemic, and further bereavements during the study.

### Characteristics of the deceased

Table 2 shows the characteristics of the deceased person for each round of surveys. Across the study, days since bereavement ranged from 1 to 812 days. The mean age of the deceased across the four timelines was either 72 or 73 and the median was 74, with a range of less than 1 year (during pregnancy) to 102 years. Over 70% of participants lost either a parent or a partner. There was an increase in the percentage of those who lost a partner across the four time-points in the study and a decrease in the percentage of those who lost a grandparent or a family member in the ‘other’ category, suggesting lower retention in the latter. A slight majority of deaths were not due to COVID- 19 and over 70% were unexpected by the bereaved respondent. Most deaths occurred in the hospital, followed by at home and in a care home. These trends were very similar across the whole study period.

**Table 2.**
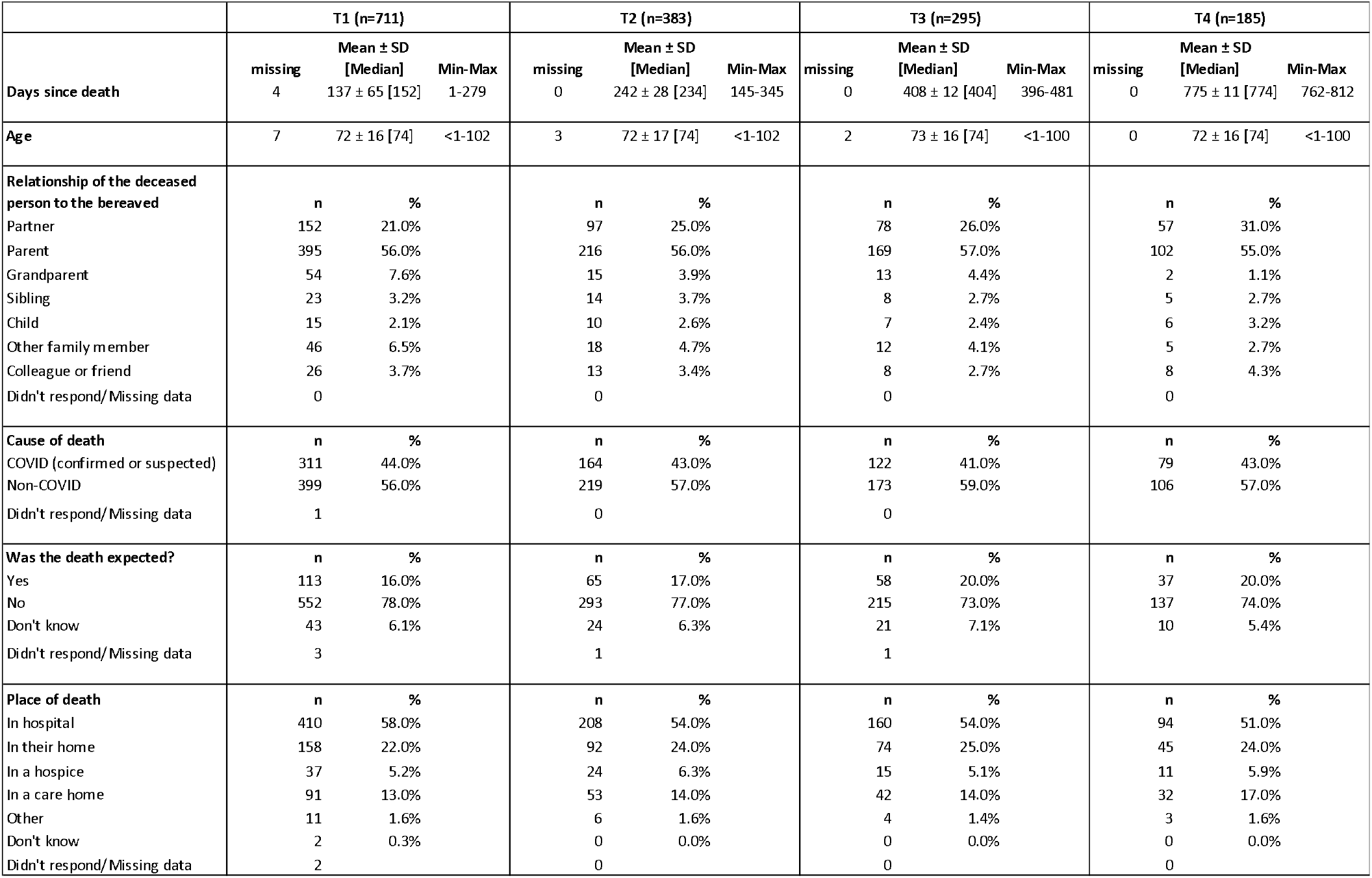
Characteristics of the deceased.

|  | T1 (n=711) |  |  | T2 (n=383) |  |  | T3 (n=295) |  |  | T4 (n=185) |  |  |
| --- | --- | --- | --- | --- | --- | --- | --- | --- | --- | --- | --- | --- |
| | missing | Mean ± SD<br>[Median] | Min-Max | missing | Mean ± SD<br>[Median] | Min-Max | missing | Mean ± SD<br>[Median] | Min-Max | missing | Mean ± SD<br>[Median] | Min-Max |
| **Days since death** | 4 | 137 ± 65 [152] | 1-279 | 0 | 242 ± 28 [234] | 145-345 | 0 | 408 ± 12 [404] | 396-481 | 0 | 775 ± 11 [774] | 762-812 |
| **Age** | 7 | 72 ± 16 [74] | <1-102 | 3 | 72 ± 17 [74] | <1-102 | 2 | 73 ± 16 [74] | <1-100 | 0 | 72 ± 16 [74] | <1-100 |
| **Relationship of the deceased person to the bereaved** | n | % |  | n | % |  | n | % |  | n | % |  |
| Partner | 152 | 21.0% |  | 97 | 25.0% |  | 78 | 26.0% |  | 57 | 31.0% |  |
| Parent | 395 | 56.0% |  | 216 | 56.0% |  | 169 | 57.0% |  | 102 | 55.0% |  |
| Grandparent | 54 | 7.6% |  | 15 | 3.9% |  | 13 | 4.4% |  | 2 | 1.1% |  |
| Sibling | 23 | 3.2% |  | 14 | 3.7% |  | 8 | 2.7% |  | 5 | 2.7% |  |
| Child | 15 | 2.1% |  | 10 | 2.6% |  | 7 | 2.4% |  | 6 | 3.2% |  |
| Other family member | 46 | 6.5% |  | 18 | 4.7% |  | 12 | 4.1% |  | 5 | 2.7% |  |
| Colleague or friend | 26 | 3.7% |  | 13 | 3.4% |  | 8 | 2.7% |  | 8 | 4.3% |  |
| Didn't respond/Missing data | 0 |  |  | 0 |  |  | 0 |  |  | 0 |  |  |
| **Cause of death** | n | % |  | n | % |  | n | % |  | n | % |  |
| COVID (confirmed or suspected) | 311 | 44.0% |  | 164 | 43.0% |  | 122 | 41.0% |  | 79 | 43.0% |  |
| Non-COVID | 399 | 56.0% |  | 219 | 57.0% |  | 173 | 59.0% |  | 106 | 57.0% |  |
| Didn't respond/Missing data | 1 |  |  | 0 |  |  | 0 |  |  |  |  |  |
| **Was the death expected?** | n | % |  | n | % |  | n | % |  | n | % |  |
| Yes | 113 | 16.0% |  | 65 | 17.0% |  | 58 | 20.0% |  | 37 | 20.0% |  |
| No | 552 | 78.0% |  | 293 | 77.0% |  | 215 | 73.0% |  | 137 | 74.0% |  |
| Don't know | 43 | 6.1% |  | 24 | 6.3% |  | 21 | 7.1% |  | 10 | 5.4% |  |
| Didn't respond/Missing data | 3 |  |  | 1 |  |  | 1 |  |  |  |  |  |
| **Place of death** | n | % |  | n | % |  | n | % |  | n | % |  |
| In hospital | 410 | 58.0% |  | 208 | 54.0% |  | 160 | 54.0% |  | 94 | 51.0% |  |
| In their home | 158 | 22.0% |  | 92 | 24.0% |  | 74 | 25.0% |  | 45 | 24.0% |  |
| In a hospice | 37 | 5.2% |  | 24 | 6.3% |  | 15 | 5.1% |  | 11 | 5.9% |  |
| In a care home | 91 | 13.0% |  | 53 | 14.0% |  | 42 | 14.0% |  | 32 | 17.0% |  |
| Other | 11 | 1.6% |  | 6 | 1.6% |  | 4 | 1.4% |  | 3 | 1.6% |  |
| Don't know | 2 | 0.3% |  | 0 | 0.0% |  | 0 | 0.0% |  | 0 | 0.0% |  |
| Didn't respond/Missing data | 2 |  |  | 0 |  |  | 0 |  |  | 0 |  |  |

### Characteristics of the experience of bereavement

Table 3 summarizes the characteristics of the experience of bereavement in terms of pandemic-related challenges before or after the death of a loved one and the support received by healthcare professionals immediately following the death. Most participants reported having had restricted funeral arrangements and limited contact with close relatives or friends (over 90% and over 80% across all time points, respectively), while a smaller majority reported a sense of isolation and loneliness (66.7% at baseline, varying by a maximum of 5.7 percentual points across all time points). Smaller majorities also reported being unable to say goodbye as they liked (63.9% at baseline, varying by a maximum of 2.9 percentual points across time points) and having limited contact with the deceased in the last days of their lives (57.8% at baseline, varying by a maximum of 1.2 percentual points across time points), while approximately half of respondents reported being unable to visit the deceased in the last days of their lives (54.3% at baseline, varying by a maximum of 8.2 percentual points across time points). Accordingly, a greater proportion of people reported facing all three challenges related to social isolation (60%, at baseline, varying by a maximum of 7 percentual points across time points) compared to the proportion of people who reported facing all three challenges related to contact prior to death (35%, at baseline, varying by a maximum of 2.6 percentual points across time points). Less than 2% of people reported not facing any of the COVID- 19 related challenges and around a quarter of respondents reported facing all six challenges. Approximately 20% reported facing three, four or five of the challenges. Over half of the participants felt little or no support by healthcare professionals and only 20% reported feeling very well supported. Changes in these proportions across time points do not suggest major bias in sample retention regarding experience of bereavement.

**Table 3.** Characteristics of the experience of bereavement regarding COVID-19 restrictions and perceived level of support offered by healthcare professionals.

|  | T1 (n=711) |  | T2 (n=383) |  | T3 (n=295) |  | T4 (n=185) |  |
| --- | --- | --- | --- | --- | --- | --- | --- | --- |
| **Contact prior to death** | **n (yes)** | **%** | **n (yes)** | **%** | **n (yes)** | **%** | **n (yes)** | **%** |
| Unable to visit them prior to their death | 386 | 54.3% | 188 | 49.0% | 136 | 46.1% | 87 | 47.0% |
| Unable to say goodbye as I would have liked | 454 | 63.9% | 238 | 62.1% | 180 | 61.0% | 114 | 61.6% |
| Limited contact with them in last days of their life | 411 | 57.8% | 222 | 58.0% | 174 | 59.0% | 107 | 57.8% |
| **Social isolation** | **n (yes)** | **%** | **n (yes)** | **%** | **n (yes)** | **%** | **n (yes)** | **%** |
| Restricted funeral arrangements | 664 | 93.4% | 363 | 94.8% | 280 | 94.9% | 176 | 95.1% |
| Limited contact with other close relatives or friends | 574 | 80.7% | 318 | 83.0% | 253 | 85.8% | 157 | 84.9% |
| Sense isolation and loneliness | 474 | 66.7% | 267 | 69.7% | 200 | 67.8% | 134 | 72.4% |
| **Number of negative experiences related to contact prior to death** | **n** | **%** | **n** | **%** | **n** | **%** | **n** | **%** |
| 0 | 147 | 21.0% | 88 | 23.0% | 74 | 25.0% | 44 | 23.8% |
| 1 | 128 | 18.0% | 69 | 18.0% | 50 | 17.0% | 34 | 18.4% |
| 2 | 185 | 26.0% | 99 | 26.0% | 73 | 25.0% | 47 | 25.4% |
| 3 | 251 | 35.0% | 127 | 33.0% | 98 | 33.0% | 60 | 32.4% |
| **Number of negative experiences related to social isolation** | **n** | **%** | **n** | **%** | **n** | **%** | **n (yes)** | **%** |
| 0 | 26 | 3.7% | 11 | 2.9% | 7 | 2.4% | 3 | 1.6% |
| 1 | 87 | 12.0% | 44 | 11.0% | 33 | 11.0% | 21 | 11.4% |
| 2 | 169 | 24.0% | 80 | 21.0% | 65 | 22.0% | 37 | 20.0% |
| 3 | 429 | 60.0% | 248 | 65.0% | 190 | 64.0% | 124 | 67.0% |
| **Total number of negative experiences** | **n** | **%** | **n** | **%** | **n** | **%** | **n** | **%** |
| 0 | 9 | 1.3% | 5 | 1.3% | 4 | 1.4% | 1 | 0.5% |
| 1 | 38 | 5.3% | 17 | 4.4% | 12 | 4.1% | 7 | 3.8% |
| 2 | 68 | 9.6% | 40 | 10.0% | 32 | 11.0% | 18 | 9.7% |
| 3 | 127 | 18.0% | 68 | 18.0% | 56 | 19.0% | 38 | 20.5% |
| 4 | 127 | 18.0% | 68 | 18.0% | 49 | 17.0% | 32 | 17.3% |
| 5 | 152 | 21.0% | 87 | 23.0% | 69 | 23.0% | 44 | 23.8% |
| 6 | 190 | 27.0% | 98 | 26.0% | 73 | 25.0% | 45 | 24.3% |
| **Felt supported by healthcare professionals** | **n** | **%** | **n** | **%** | **n** | **%** | **n** | **%** |
| Not at all supported | 252 | 35.0% | 138 | 36.0% | 101 | 34.0% | 68 | 36.8% |
| A little bit supported | 139 | 20.0% | 70 | 18.0% | 62 | 21.0% | 44 | 23.8% |
| Fairly well supported | 105 | 15.0% | 61 | 16.0% | 44 | 15.0% | 23 | 12.4% |
| Very well supported | 95 | 13.0% | 61 | 16.0% | 50 | 17.0% | 30 | 16.2% |
| Not relevant (not next of kin) | 120 | 17.0% | 53 | 14.0% | 38 | 13.0% | 20 | 10.8% |
| Didn't respond/Missing data | 0 |  | 0 |  | 0 |  | 0 |  |

### Grief and social support outcomes

Table 4 summarizes the participants’ outcome measures across the study.

**Table 4.**
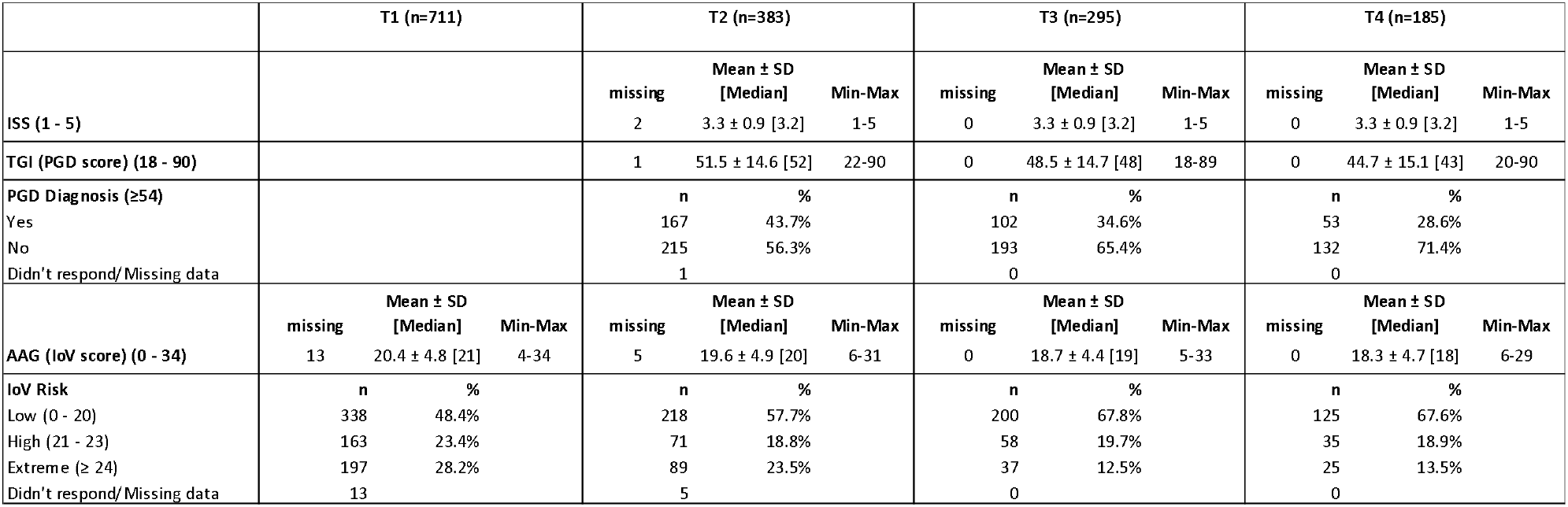
Summary of participants’ grief and support outcomes across the study.

|  | T1 (n=711) |  |  | T2 (n=383) |  |  | T3 (n=295) |  |  | T4 (n=185) |  |  |
| --- | --- | --- | --- | --- | --- | --- | --- | --- | --- | --- | --- | --- |
| | | | | Mean ± SD<br>[Median] Min-Max | | | Mean ± SD<br>[Median] Min-Max | | | Mean ± SD<br>[Median] Min-Max | | |
| ISS (1 - 5) | | | | missing<br>2 | 3.3 ± 0.9 [3.2] | 1-5 | missing<br>0 | 3.3 ± 0.9 [3.2] | 1-5 | missing<br>0 | 3.3 ± 0.9 [3.2] | 1-5 |
| TGI (PGD score) (18 - 90) | | | | 1 | 51.5 ± 14.6 [52] | 22-90 | 0 | 48.5 ± 14.7 [48] | 18-89 | 0 | 44.7 ± 15.1 [43] | 20-90 |
| PGD Diagnosis ($\geq 54$) | | | | n | % | | n | % | | n | % | |
| Yes |  |  |  | 167 | 43.7% |  | 102 | 34.6% |  | 53 | 28.6% |  |
| No |  |  |  | 215 | 56.3% |  | 193 | 65.4% |  | 132 | 71.4% |  |
| Didn't respond/Missing data |  |  |  | 1 |  |  | 0 |  |  | 0 |  |  |
| | missing | Mean ± SD<br>[Median] | Min-Max | missing | Mean ± SD<br>[Median] | Min-Max | missing | Mean ± SD<br>[Median] | Min-Max | missing | Mean ± SD<br>[Median] | Min-Max |
| AAG (ioV score) (0 - 34) | 13 | 20.4 ± 4.8 [21] | 4-34 | 5 | 19.6 ± 4.9 [20] | 6-31 | 0 | 18.7 ± 4.4 [19] | 5-33 | 0 | 18.3 ± 4.7 [18] | 6-29 |
| ioV Risk | n | % |  | n | % |  | n | % |  | n | % |  |
| Low (0 - 20) | 338 | 48.4% |  | 218 | 57.7% |  | 200 | 67.8% |  | 125 | 67.6% |  |
| High (21 - 23) | 163 | 23.4% |  | 71 | 18.8% |  | 58 | 19.7% |  | 35 | 18.9% |  |
| Extreme ($\geq 24$) | 197 | 28.2% | | 89 | 23.5% | | 37 | 12.5% | | 25 | 13.5% | |
| Didn't respond/Missing data | 13 |  |  | 5 |  |  | 0 |  |  | 0 |  |  |

TGI: Mean TGI (PGD) score at T2 was 51.5. This decreased steadily across survey time points, dropping to 48.5 at T3 and 44.7 at T4. At T2 43.7% met the threshold for indicated PGD (≥54), dropping to 34.6% at T3 (c.13 months post-bereavement) and 28.6% at final follow up, c. 25 months post-bereavement.

AAG: Mean IOV (grief vulnerability) score at baseline was 20.4, decreasing slightly but steadily to 18.7 at T3 and 18.3 at 24. At baseline 48.4% exhibited low levels of vulnerability (i.e., 0 ≤ IOV ≤ 20); 23.4% exhibited high levels (i.e., 21 ≤ IOV ≤ 23), and 28.2% exhibited severe levels (i.e., IOV ≥ 24). By T4 67% exhibited low levels of vulnerability, 18.9% exhibited high levels and 13.5% demonstrated severe levels.

ISS: Social support scores were stable and did not change across survey time points. Mean SSI score at T2,T3,T4 was 3.3, see table 4. This could be interpreted as feeling ‘fairly well supported’, with a score of 5 meaning very well and a score of 1 meaning ‘not at all’.

### Factors associated with levels of prolonged grief, social support and vulnerability in grief

Table 5 shows the effect sizes for each individual variable from the single models on PGD, ISS and IOV and the R values for the models containing each set of variables. Across all the analyses, characteristics of the deceased were generally and consistently the best predictors of all three indices: PGD, ISS and IOV. For PGD and IOV, relationship with deceased, followed by place of death, showed the largest effect sizes across all variables, whilst for ISS, the largest effects were from feeling supported by health care professionals following the death and ethnicity, only then followed by place of death and relationship with deceased. Ethnicity also showed a large effect on PGD, with white respondents showing worse grief outcomes compared to minoritized ethnic respondents, but not on IOV. Qualifications showed large effects for PGD and IOV, with those from lower education levels showing worse grief outcomes, but not for ISS. All items related to contact prior to death showed small effects for all three indices, while items for social isolation showed large and very large effects for PGD and IOV; specifically sense of isolation and loneliness had a very large effect on PGD and a large effect on IOV, while restricted funeral arrangements had a large effect on PGD but a medium effect on IOV. Days since death had a larger effect on PGD than IOV or ISS; even though the effect of the slope is not very large even for PGD, it represents a considerable change in PGD over an extended period of time (e.g. an approximate reduction of 4 scale points in TGI for each 5 months).

**Table 5.**
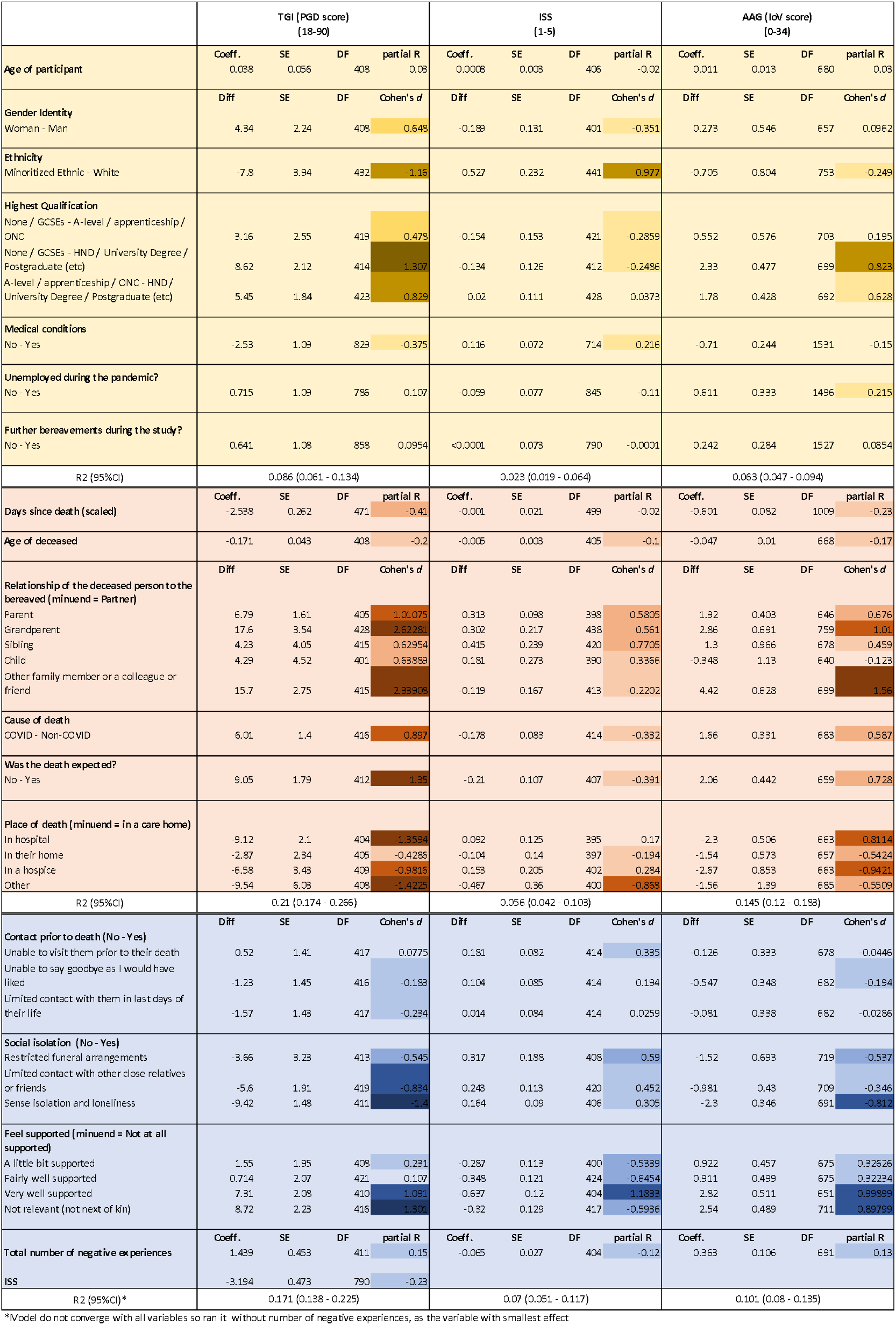
Results of the mixed models with participant ID as random term, days since bereavement as co-variate and each of the predictors individually. The table is split by groups of variables: characteristics of the participants in yellow; characteristics of the deceased in salmon and characteristics of the experience of bereavement in blue. R-squared are presented for the models containing each group of variables. For the partial R and the Cohen’s d, the intensity of the shading reflects the strength of the effect.

Table 6 shows the model outputs and Figures 1 to 3 show the model predictions for PGD, ISS and IOV for variables that had the largest effect sizes.

**Figure 1.**
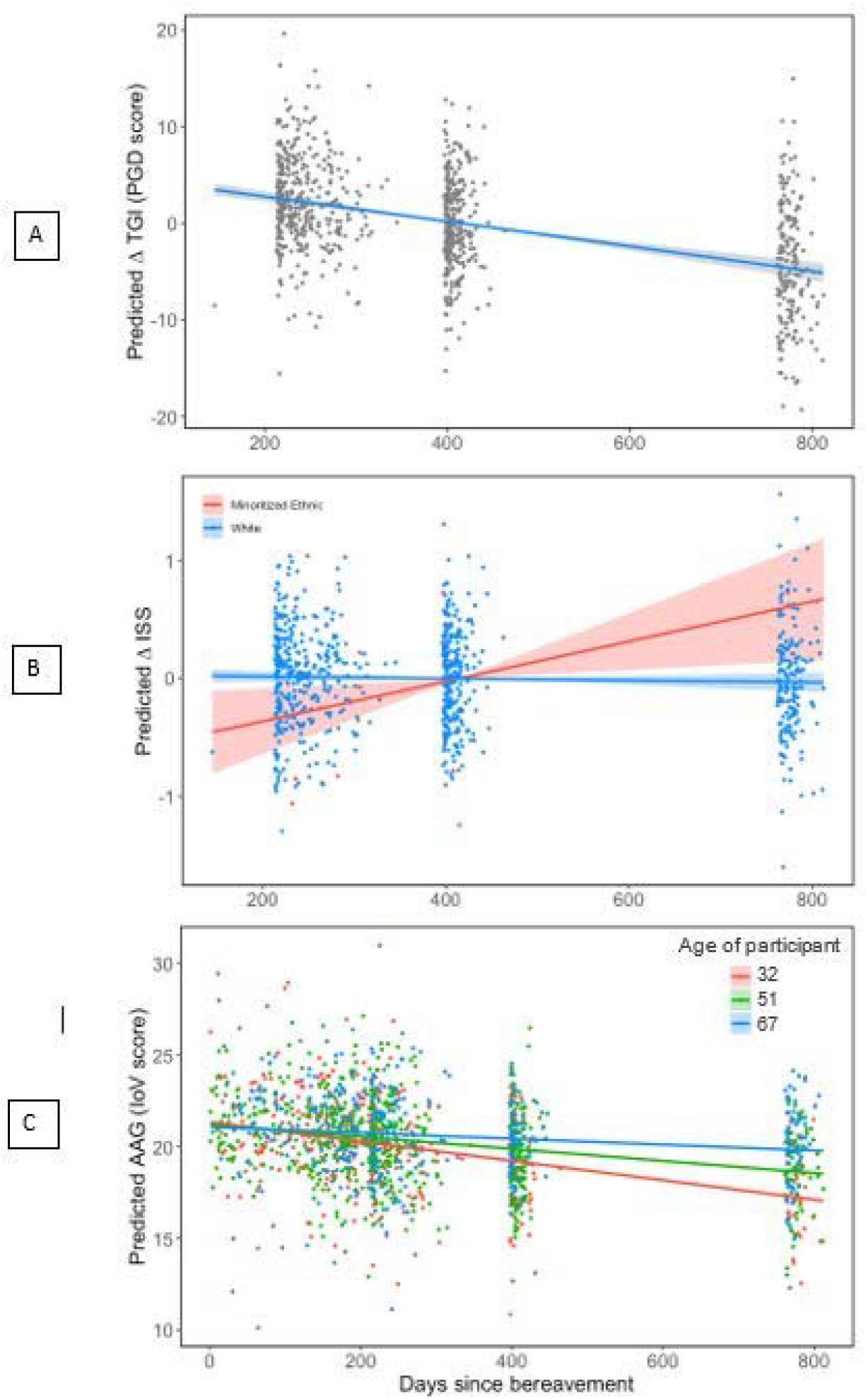
Predicted temporal patterns of RGD scores (A), ISS (B) and IoV (C) scores regarding days since bereavement. ISS and IoV score showed significant interactions between days since bereavement and respectively, ethnicity and age of participant, which are depicted here. Note that PGD scores and ISS are plotted as contrasts, showing the predicted average change in PGD score and ISS across all groups of participants, while IoV is depicted as predicted scores for the first, second and third quartiles or participants ages while keeping all other variables constant (set for the group with larger sample size); predicted scores were plotted instead of contrasts because the interaction was more visually easily interpreted this way.

**Table 6.** Outcomes of the General Linear Mixed models for TGI score, ISS, and AAG score with participant ID as random term, days since bereavement as covariate and all the predictors in combination (excluding those with very small effects for each given model that were necessary to remove for the model to fit appropriately - total number of negative experiences was included in the starting models but was removed from all the models due to small effects).

| *Predictors* | **TGI (PGD score)** |  |  | **ISS** |  |  | **AAG (IoV score)** |  |  |
| --- | --- | --- | --- | --- | --- | --- | --- | --- | --- |
|  | *Estimates* | *CI* | *p* | *Estimates* | *CI* | *p* | *Estimates* | *CI* | *p* |
| (Intercept) | 68.31 | 54.26 – 82.37 | **<0.001** | 3.91 | 2.95 – 4.87 | **<0.001** | 22.97 | 20.07 – 25.87 | **<0.001** |
| Days (scaled) | -2.69 | -3.21 – -2.17 | **<0.001** | 0.35 | 0.08 – 0.63 | **0.011** | -1.88 | -2.57 – -1.19 | **<0.001** |
| Gender [Male] | -1.86 | -5.84 – 2.13 | 0.361 | 0.14 | -0.13 – 0.42 | 0.298 |  |  |  |
| Ethnicity grouped [White] | 4.9 | -2.40 – 12.21 | 0.188 | -0.18 | -0.70 – 0.33 | 0.487 |  |  |  |
| Medical conditions [Yes] | 2.17 | 0.21 – 4.13 | **0.03** | -0.12 | -0.26 – 0.02 | 0.102 |  |  |  |
| Unemployed [Yes] |  |  |  | 0.01 | -0.14 – 0.17 | 0.854 |  |  |  |
| Highest qualification grouped [A-level / apprenticeship / ONC] | -1.79 | -6.25 – 2.67 | 0.432 | 0.07 | -0.24 – 0.37 | 0.663 | -0.08 | -1.14 – 0.98 | 0.886 |
| Highest qualification grouped [HND / University Degree / Postgraduate (etc)] | -5.19 | -9.01 – -1.38 | **0.008** | 0.03 | -0.24 – 0.29 | 0.851 | -1.27 | -2.19 – -0.35 | 0.007 |
| Age of participant |  |  |  | -0.01 | -0.02 – 0.01 | 0.372 | 0.02 | -0.02 – 0.06 | 0.27 |
| Relationship [Parent] | -4.18 | -7.49 – -0.87 | **0.013** | -0.38 | -0.72 – -0.03 | **0.031** | -0.57 | -1.75 – 0.60 | 0.34 |
| Relationship [Sibling] | -3.58 | -10.87 – 3.70 | 0.335 | -0.17 | -0.66 – 0.32 | 0.499 | -1.73 | -3.62 – 0.16 | 0.072 |
| Relationship [Child] | -11.42 | -20.58 – -2.25 | **0.015** | -0.29 | -0.93 – 0.36 | 0.383 | -1.32 | -3.66 – 1.01 | 0.267 |
| Relationship [Grandparent] | -10.03 | -17.84 – -2.22 | **0.012** | -0.44 | -1.19 – 0.31 | 0.252 | -0.47 | -2.74 – 1.80 | 0.683 |
| Relationship [Other] | -10.52 | -16.37 – -4.68 | **<0.001** | 0.16 | -0.26 – 0.58 | 0.465 | -2.84 | -4.26 – -1.42 | **<0.001** |
| Death expected [Yes] | -6.39 | -9.96 – -2.81 | **<0.001** | 0.04 | -0.20 – 0.28 | 0.737 | -1.42 | -2.32 – -0.51 | **0.002** |
| Place of death [In a hospice] | 5.5 | -1.02 – 12.02 | 0.098 | -0.44 | -0.88 – -0.00 | **0.047** | 3.24 | 1.58 – 4.89 | **<0.001** |
| Place of death [In hospital] | 4.77 | 0.91 – 8.64 | **0.016** | -0.16 | -0.42 – 0.10 | 0.241 | 1.42 | 0.45 – 2.39 | **0.004** |
| Place of death [In their home] | 3.45 | -1.23 – 8.14 | 0.148 | 0 | -0.31 – 0.32 | 0.991 | 2 | 0.82 – 3.18 | **0.001** |
| Place of death [Other] | 8.46 | -3.11 – 20.04 | 0.152 | 0.48 | -0.29 – 1.26 | 0.221 | 0.95 | -1.73 – 3.62 | 0.487 |
| Cause of death [Non-Covid] | -1.32 | -4.42 – 1.78 | 0.405 | 0 | -0.21 – 0.21 | 0.978 | -0.93 | -1.70 – -0.17 | **0.016** |
| Age of deceased person | -0.11 | -0.21 – -0.00 | **0.043** | 0 | -0.01 – 0.01 | 0.769 | -0.05 | -0.08 – -0.01 | **0.011** |
| Limited contact last days [yes] | 1.44 | -1.36 – 4.25 | 0.313 | 0.14 | -0.06 – 0.33 | 0.17 | -0.24 | -0.95 – 0.47 | 0.506 |
| Restricted funeral [yes] | -1.7 | -7.54 – 4.14 | 0.567 | -0.18 | -0.57 – 0.22 | 0.385 | -0.02 | -1.37 – 1.33 | 0.974 |
| Isolation [yes] | 5.86 | 2.75 – 8.97 | **<0.001** | -0.05 | -0.26 – 0.17 | 0.664 | 1.4 | 0.64 – 2.16 | **<0.001** |
| Limited contact relatives [yes] | 0.46 | -3.26 – 4.17 | 0.809 | -0.11 | -0.36 – 0.14 | 0.372 | -0.18 | -1.06 – 0.69 | 0.681 |
| Able to visit [yes] | -2.29 | -5.17 – 0.58 | 0.118 | -0.13 | -0.33 – 0.06 | 0.181 | -0.23 | -0.95 – 0.49 | 0.528 |
| Able to say goodbye [yes] | -1.43 | -4.71 – 1.84 | 0.39 | 0.14 | -0.08 – 0.36 | 0.221 | 0.03 | -0.78 – 0.84 | 0.944 |
| Feel supported [A little bit supported] | -0.55 | -3.92 – 2.82 | 0.749 | 0.27 | 0.04 – 0.50 | **0.02** | -0.8 | -1.64 – 0.03 | 0.06 |
| Feel supported [Fairly well supported] | 0.54 | -3.14 – 4.22 | 0.774 | 0.32 | 0.07 – 0.57 | **0.011** | -0.37 | -1.33 – 0.58 | 0.443 |
| Feel supported [Not relevant to my situation] | -2.06 | -6.80 – 2.67 | 0.392 | 0.12 | -0.19 – 0.44 | 0.446 | -1.04 | -2.12 – 0.05 | 0.062 |
| Feel supported [Very well supported] | -4.23 | -8.15 – -0.31 | **0.034** | 0.6 | 0.33 – 0.86 | **<0.001** | -2.46 | -3.46 – -1.47 | **<0.001** |
| ISS | -3.12 | -4.04 – -2.21 | **<0.001** |  |  |  |  |  |  |
| Days (scaled) × Ethnicity grouped [White] |  |  |  | -0.37 | -0.64 – -0.10 | **0.008** |  |  |  |
| Days (scaled) × Age of participant |  |  |  |  |  |  | 0.02 | 0.01 – 0.04 | **<0.001** |
| **Random Effects** |  |  |  |  |  |  |  |  |  |
| $\sigma^2$ | 43.31 | | | 0.28 | | | 7.91 | | |
| $\tau_{00\_PID\_shortened}$ | 119.58 | | | 0.48 | | | 10.7 | | |
| ICC | 0.73 |  |  | 0.63 |  |  | 0.57 |  |  |
| N $PID\_shortened$ | 411 | | | 402 | | | 672 | | |
| Observations | 842 |  |  | 821 |  |  | 1510 |  |  |
| Marginal $R^2$ / Conditional $R^2$ | 0.311 / 0.817 | | | 0.119 / 0.674 | | | 0.206 / 0.662 | | |

### Effect of time

Figure 1 shows trends of PGD, ISS and IOV in relation to time since bereavement. There was a general tendency for PGD and IOV to improve with time, which was more noticeable for PGD than for IOV. Furthermore, there was an interaction between days since bereavement and age of participant for IOV, with younger participants improving more through time compared to older participants. The relationship between days since bereavement and ISS is less strong with a slight tendency for ISS to improve with time, although this is mainly driven by those from minoritized ethnic groups, with white participants showing no change in ISS through time. Although only the linear trend was fitted for simplicity, visual analysis showed that the sharpest decline in grief scores seem to occur between 6 months to a year since bereavement.

These were general patterns averaged across participants, but it was noticeable that for all indices of bereavement different participants would show different patterns, with some improving through time, some worsening through time, some showing oscillations but no real trend and some showing no change (Figure 2).

**Figure 2.**
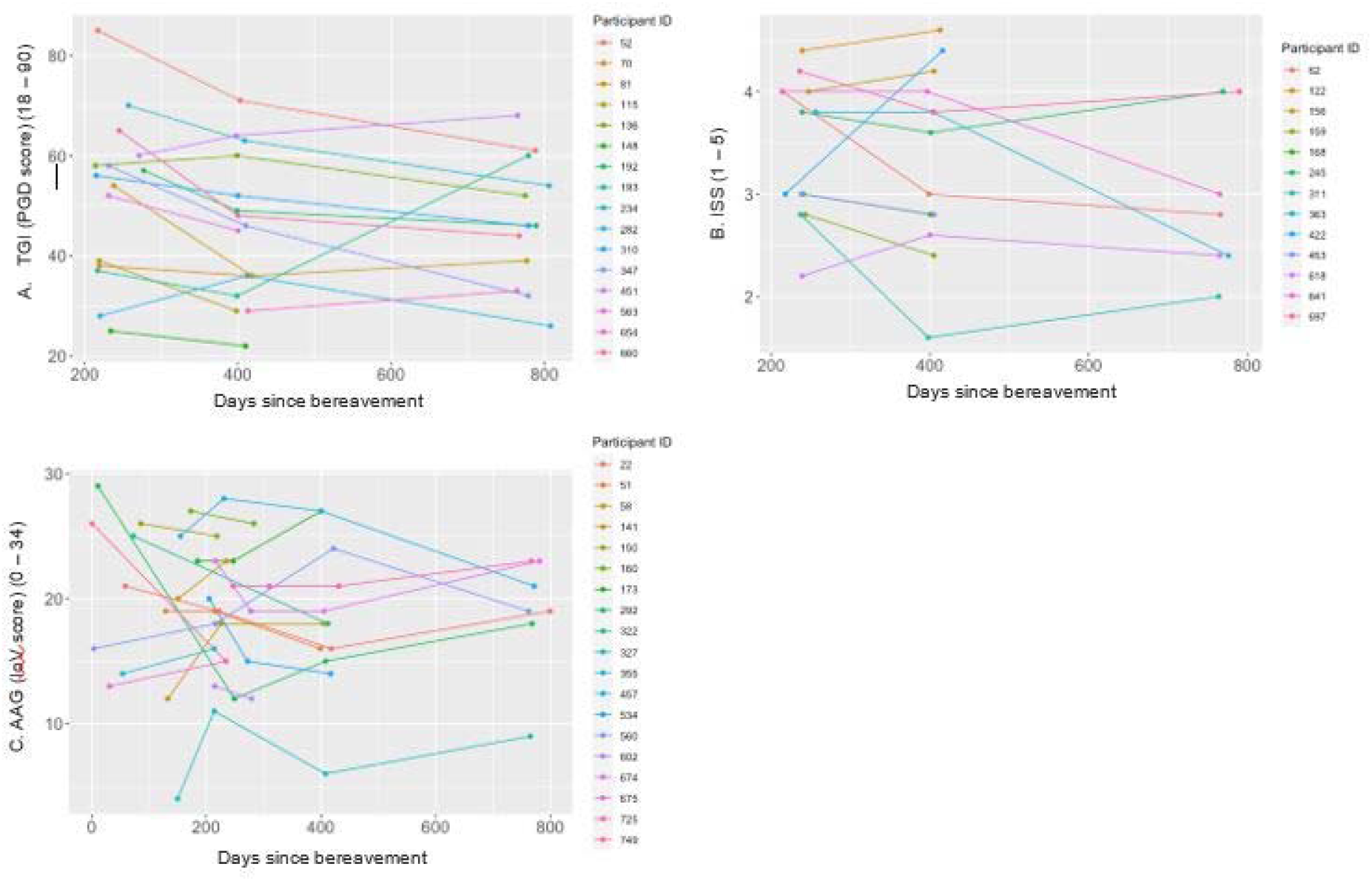
Changes in PGD scores (A), ISS (B) and IoV(C) scores through time for different randomly generated selections of study participants

### Effect of experience of bereavement

The results from the full models examining the effects of the experiences of bereavement while controlling for characteristics of the participant and the bereaved, showed that sense of isolation and feeling supported by healthcare professionals were the most important experiences in predicting PGD and IOV scores, while only the latter was an important predictor of ISS. Those who felt very well supported by healthcare professionals showed better grief and support outcomes compared to all other groups. ISS score was also an important predictor of PGD; In the same model but without ISS, feeling supported by healthcare professionals showed a much larger effect on PGD than when ISS was included, which suggests that ISS has a mediating effect between feeling supported and PGD levels, while sense of isolation has both a direct and indirect effect on PGD. These patterns are depicted in Figure 3.

**Figure 3.**
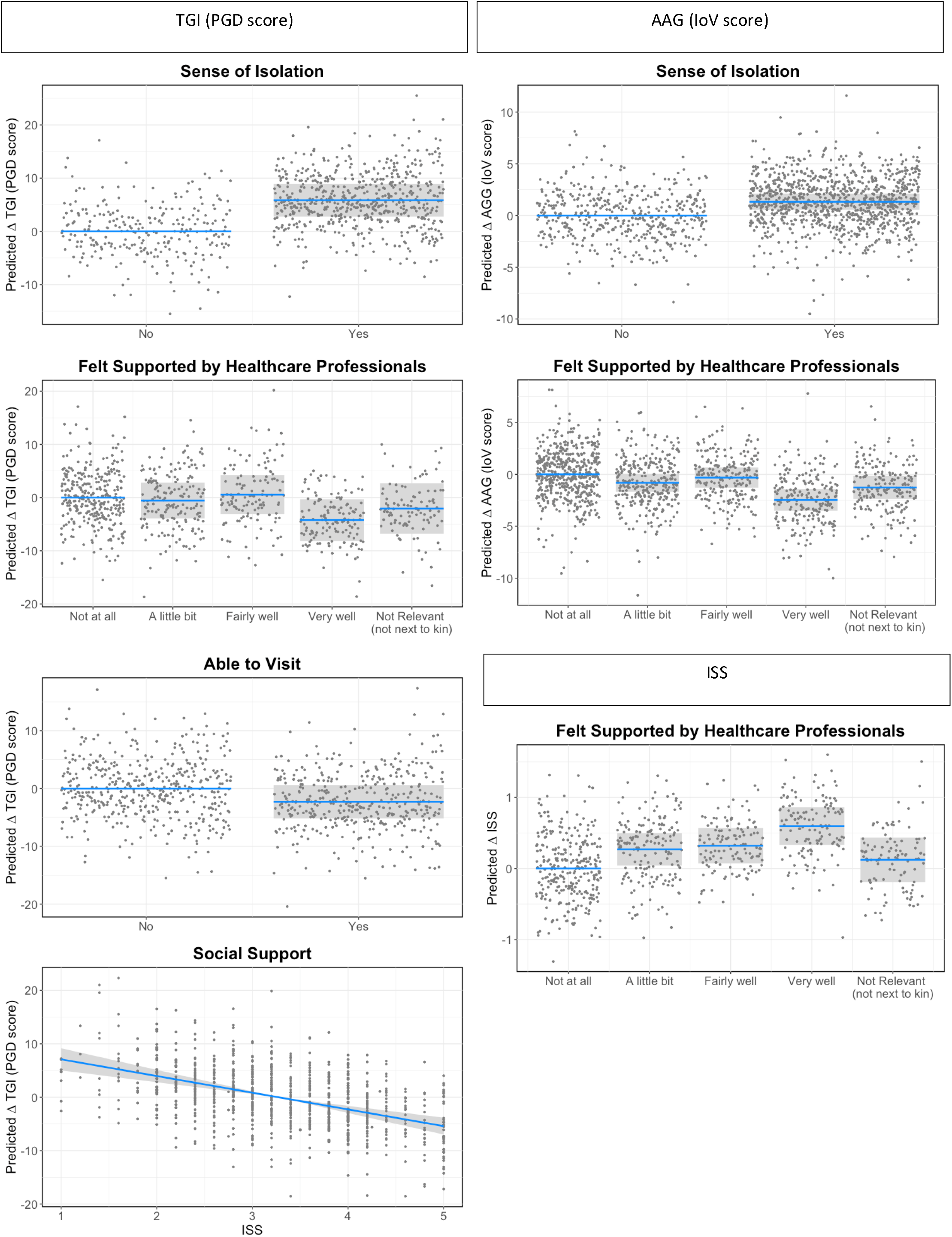
Predicted changes (statistical contrasts) in PGD scores, ISS and IoV scores across different experiences of bereavement identified in the statistical models as having the strongest effect sizes for each outcome. Shaded areas represent 95% confidence intervals for the change in PGD, ISS or IoV score in relation to the reference category. Note that PGD scores range from 18 to 90, IoV from 0 to 34 and ISS from 1 to 5, and hence the same change in PGD and IoV represents an approximate twofold change in magnitude for IoV compared to PGD, while a change in ISS represents an approximate sevenfold change in magnitude compared to IoV and 14-fold change in magnitude compared to PGD.

Despite representing a relatively modest effect, being unable to visit a loved one prior to death had an increased effect on PGD once all other factors were accounted for (an estimated average difference of 2.3 scale points in PGD level). Restricted funeral arrangements showed a much smaller effect for IOV when other variables were controlled for than in the single model. Number of negative experiences during bereavement showed very low effect sizes across all three indices, once other variables were accounted for.

### Effect of the characteristics of the participant and the deceased

Place of death and relationship with deceased remained strongest predictors of grief and support outcomes, although the effect of relationship with deceased was generally smaller when other factors were accounted for, most noticeable for IOV. Specifically, the estimated differences in IOV between losing a partner or a parent or grandparent were much smaller than when the variable was considered in the single model. Those bereaved of a partner showed higher PGD scores and slightly higher IOV scores compared to other groups. Those bereaved of a child showed smaller PGD scores than all other groups and also relatively small IOV scores but the highest ISS scores. Those bereaved of a parent or grandparent had lower ISS than those whose partner died, with bereaved partners’ ISS scores similar to those bereaved of a more distant relative.

There were generally worse outcomes for those who had a loved one dying in a hospice and best for those who had a loved one dying in a care home, with PGD scores also high for those on the ‘other’ category. ISS scores were lowest for deaths in hospices and highest for the ‘other’ category.

An unexpected death had a negative effect in both PGD and IOV, while cause of death only had a meaningful influence on IOV – those who lost someone to COVID-19 had slightly worse outcomes, while age of deceased only had a meaningful influence on PGD, with worse outcomes for bereavements of younger people. Although only the linear trend was fitted for simplicity, visual analysis of the relationship between age of deceased and PGD showed a sharper decrease from the age of 70 years (this was also the case for IOV and ISS scores also showed a slightly more pronounced decrease from this point). These patterns are shown in Figure 4.

**Figure 4.**
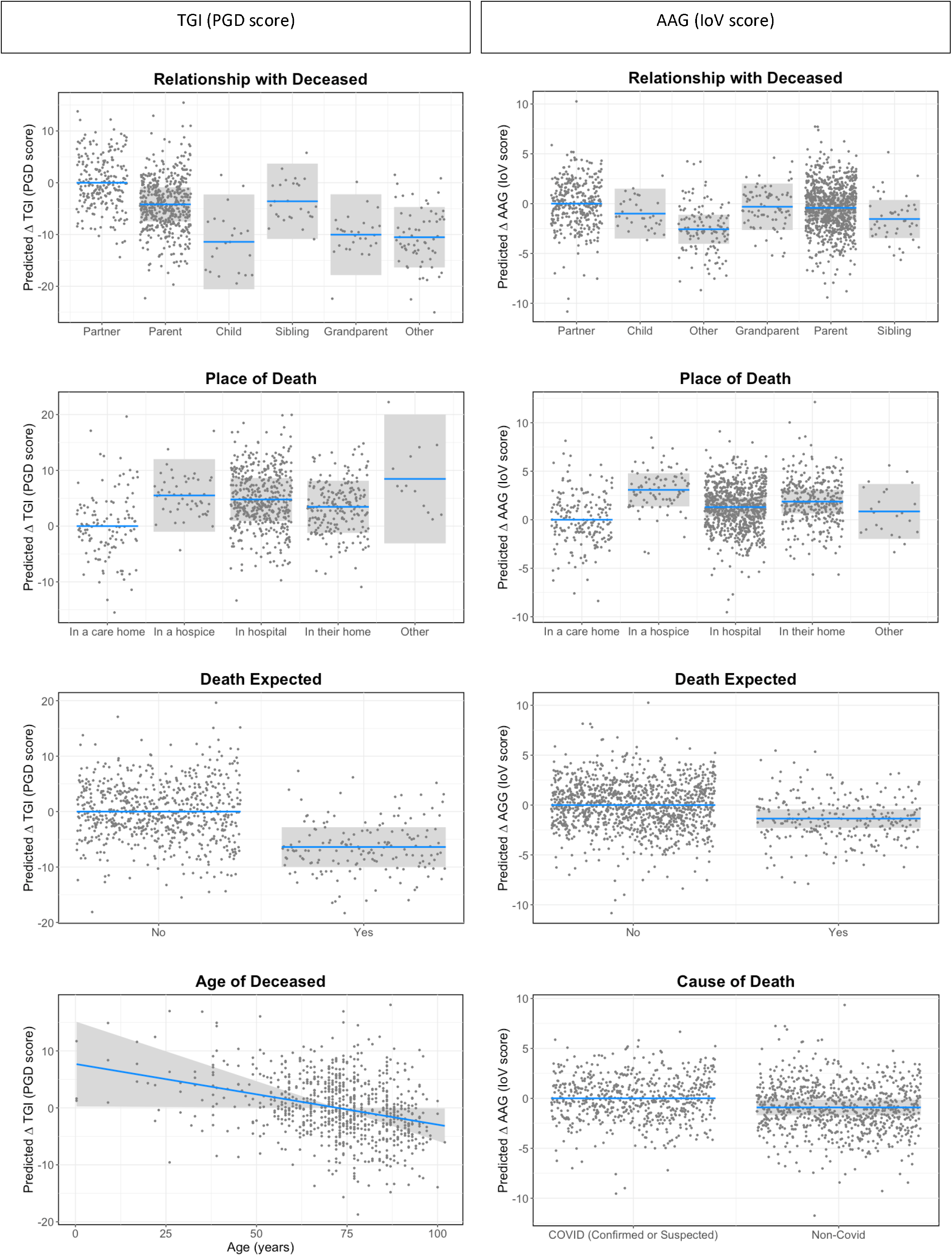
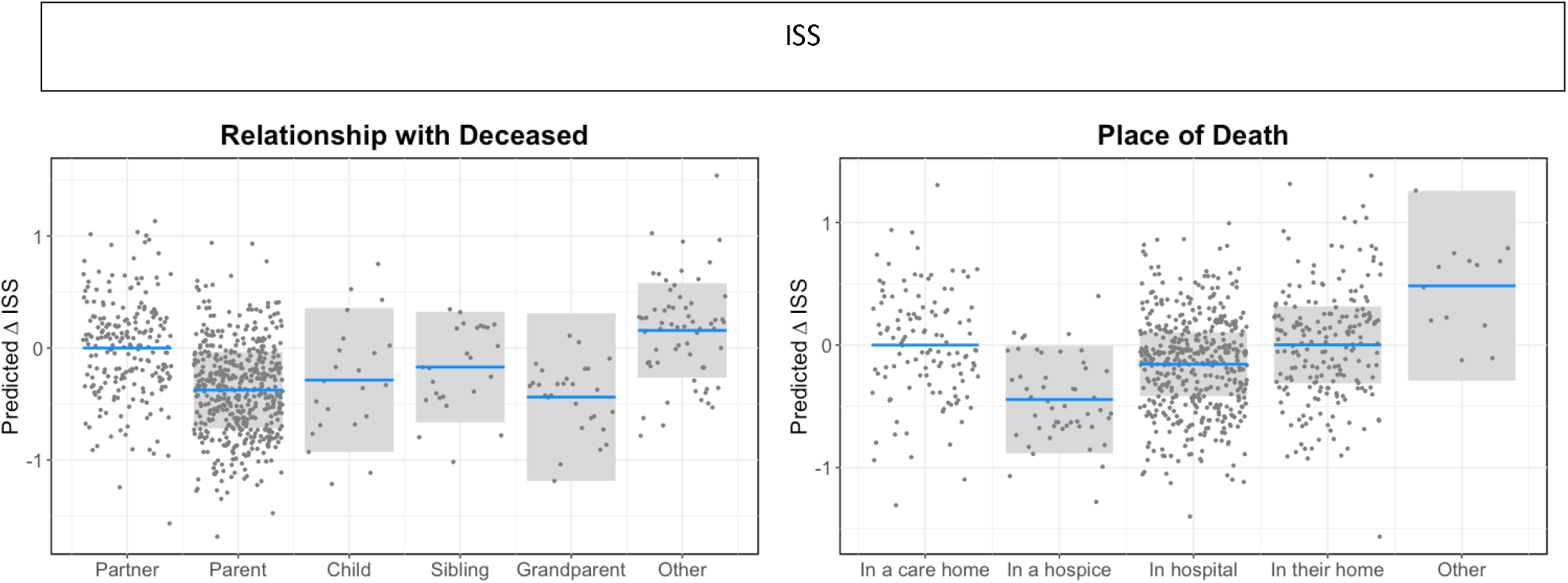
Predicted changes (statistical contrasts) in PGD and IoV scores in relation to characteristics of the bereaved and circumstances of death identified in the statistical models as having the strongest effect sizes for each outcome. Shaded areas represent 95% confidence intervals for the change in PGD, ISS or IoV score in relation to the reference category. Note that PGD scores range from 18 to 90, IoV from 0 to 34 and ISS from 1 to 5, and hence the same change in PGD and IoV represents an approximate twofold change in magnitude for IoV compared to PGD, while a change in ISS represents an approximate sevenfold change in magnitude compared to IoV and 14-fold change in magnitude compared to PGD.

Qualifications also showed important effects for PGD and IOV, with those from higher education levels showing better outcomes, and having medical conditions showed a negative effect for PGD. Ethnicity showed an estimated average difference of around 5 scale points in PGD level (with higher PGD scores for white group), but due to the small sample size and large variability in the minoritized ethnic group and potentially its relatively high association with ISS, this variable has an overall small effect size on PGD when controlling for other variables. These patterns are shown in Figure 5.

**Figure 5.**
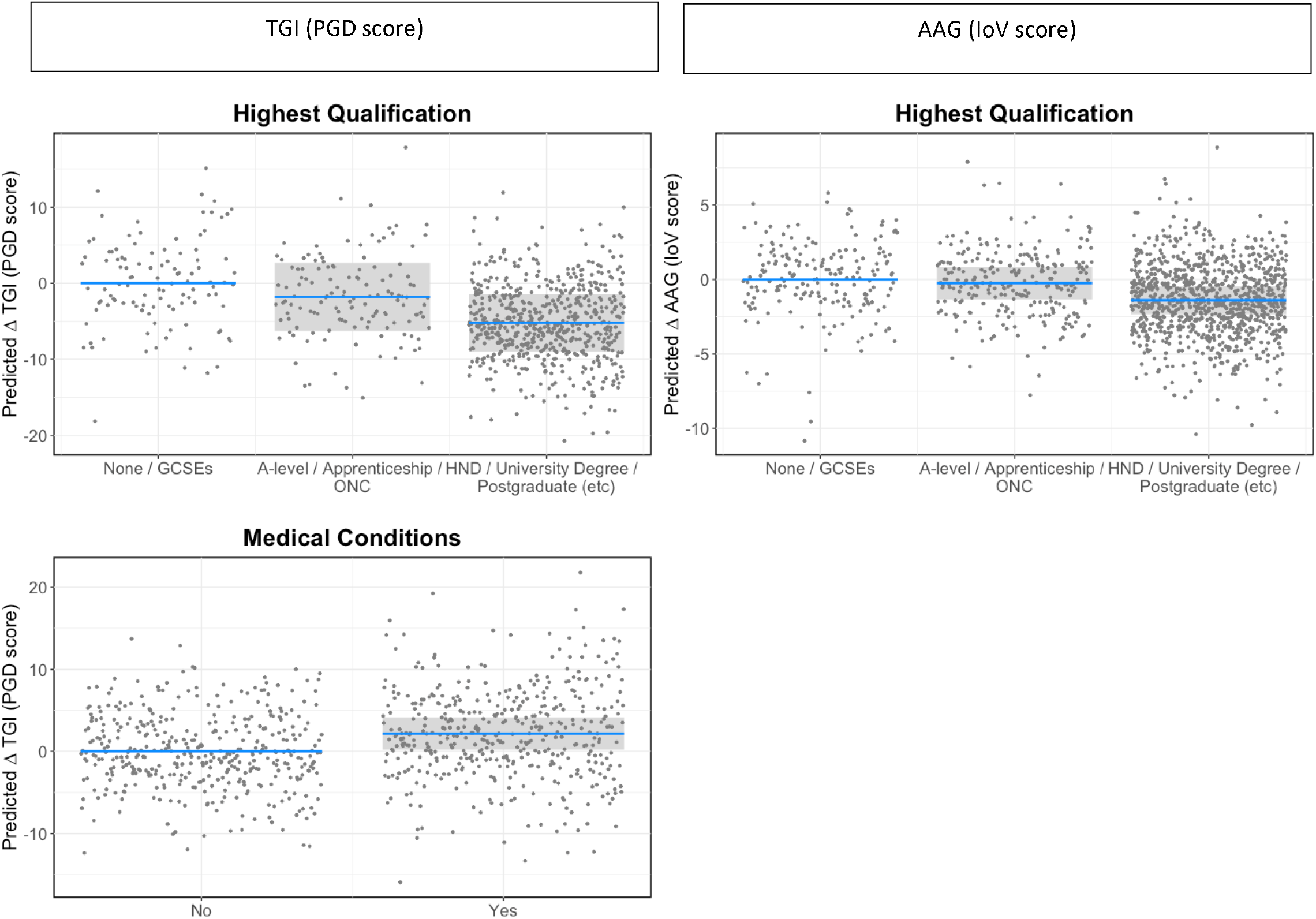
Predicted changes (statistical contrasts) in PGD and loV scores in relation to characteristics of the participants identified in the statistical models as having the strongest effect sizes for each outcome. Shaded areas represent 95% confidence intervals for the change in PGD or loV score in relation to the reference category. Note that PGD scores range from 18 to 90 and loV from 0 to 34, hence the same change in PGD and loV represents an approximate twofold change in magnitude for loV compared to PGD.

Age of participant, further bereavements during the study, and whether people were unemployed during the pandemic showed the lowest effect sizes across all three indices, once other variables were accounted for.

## DISCUSSION

This analysis represents the only longitudinal examination of COVID-19 pandemic grief outcomes that we are aware of to date, with a focus on PGD as our primary outcome. In a sample of people bereaved during the first nine months of the COVID-19 pandemic in the UK, we found decreasing but relatively high levels of indicated PGD at c. 8 (44%), 13 (34%) and 25 (27%) months post-bereavement. Factors most strongly associated with PGD scores were those relating to the person who died, in terms of their relationship to the participant, where they died and whether the death was expected. Large effects relating to support were also observed, specifically isolation and loneliness around the time of bereavement and perceived social support over time, with support from healthcare professionals immediately following the death also a factor. Level of education and existence of medical conditions were the main participant characteristics found to have an effect. These findings have important implications for bereavement policy, provision and practice in the current COVID-recovery and post-pandemic period (e.g. strengthening of specialist and social support) and in preparedness for future pandemics and mass bereavement events (e.g. infection control measures and rapid support responses).

### Grief levels

We found relatively high levels of indicated PGD and grief vulnerability (IOV) overall, and across time. As in other studies, time since death was negatively associated with PGD scores (6,34), and to a lesser extent levels of grief vulnerability (IoV). However, the proportions of people meeting the threshold for indicative PGD remained higher than would be expected in non-pandemic times, with 34% at c. 13 months bereaved and 27% at c. 25 months. Public health models of bereavement (7,11) suggest that in non-pandemic times, around 10% of bereaved people are at high risk of PGD and may need professional mental health support, and a further 30% are at moderate risk and may need some additional support e.g. via peer support groups. These estimates were confirmed in a 2015 Australian survey, which in a sample of people on average 14 months bereaved, identified 6.4% at high-risk of PGD, 35.2% at moderate risk and the remaining 58.4% at low risk (7). Although our sample is limited by its self-selecting design and is not representative, our findings would nonetheless appear to support predictions that grief disorder prevalence would rise because of the pandemic (2–4), providing longer-term evidence that is consistent with the results of earlier cross-sectional studies with more recently bereaved populations (14–20).

We now summarise the factors associated with poorer grief outcomes and consider which of these factors might explain the apparently higher levels of indicated PGD that we observed.

### The person who died

We found that, over time, relationship to the deceased continued to be strongly associated with PGD and IoV scores, as in our baseline analysis (23). People who lost a partner, child or sibling showed higher levels of grief compared with bereavements of more distant relatives/friends, although these effects became relatively less important once other factors were controlled for, especially in relation to the IoV. This effect was influenced by perceived social support in the PGD model, with the poorer social support experienced by siblings a possible factor contributing to their relatively worse PGD scores, and the better support experienced by bereaved spouses/partners seemingly buffering the effects of this loss, despite their overall poorer PGD scores. These varying levels of support also demonstrate greater perceived lack of understanding and empathy within social networks in relation to deaths of siblings, parents and grandparents, or greater reluctance to seek or ask for help amongst those experiencing these types of bereavement, as also indicated in our qualitative findings (21).

Age of the deceased had a small but significant effect on PGD scores (although not IoV scores, unlike our baseline analysis, 23), with younger age associated with higher PGD scores. These associations between relationship with and age of the deceased are consistent with pre-pandemic studies (7,10,30,51) and some studies of pandemic bereavement (6, 12–14,29). Although our sample is not directly comparable to Aoun et als (2015) study (7), which provided empirical data for the proportions of low, moderate and high-risk groups reported above, comparisons of these participant characteristics (relationship with/age of deceased) would suggest that these two factors, albeit important predictors of grief severity, cannot explain the higher grief levels observed in our pandemic study. Mean age of deceased was very similar (72 vs. 75), whilst the Australian study included higher proportions of people who had lost spouses and children, and a similar proportion of people whose siblings had died.

### Cause, expectedness and place of death

While worse outcomes have been identified for people bereaved by COVID-19 compared with pre-pandemic general bereaved populations (12-14,17), or ‘natural’ causes of death before and during the pandemic (18–20), our analysis found no effect on PGD scores for cause of death (COVID-19 vs. non-COVID-19) when other factors were controlled for (15), but a small and significant effect on IoV scores (unlike our baseline analysis, 23). However, it should also be noted that several of these factors/co-variates were associated with both COVID-19 deaths (22) and worse grief outcomes (discussed below e.g. reduced support from healthcare professionals, loneliness and isolation). As in other studies, unexpected deaths were found to have a significant negative effect on PGD and IOV (19,20,50). The fact that a much larger group of participants reported that the death was not expected (78%) than those bereaved by COVID-19 (45%), would suggest that that this aspect of a death, which likely increased in relation to both COVID and non-COVID deaths during the pandemic (e.g. due to cancer treatment delays, disruption to services, 52,53) may be an explanatory factor for elevated PGD levels during the pandemic.

As in our baseline analysis (23), place of death was also found to be strongly associated with levels of grief (IoV and PGD). Despite care-home deaths being associated with worse experiences of end-of-life care and visiting restrictions (22), with the troubling consequences of prolonged periods of separation also described qualitatively (24), grief outcomes (IoV and PGD) were better when a death occurred in a care home compared with other settings (controlling for aspects of end-of-life experience, e.g. support from healthcare professionals after the death, being able to visit). This relationship may be due to anticipatory grief work, e.g. in the context of dementia diagnoses, and reflect the more ‘expected’ nature of some of these deaths. A US study similarly found that deaths from dementia during the pandemic were negatively associated with probable PGD compared with deaths from other causes (18). The finding that those reporting ‘Other’ places of death had worst PGD scores (although a very small group-size), followed by hospice and hospital deaths might reflect the consequences of especially traumatic, sudden deaths (e.g. accidents) (7,19,20), the more difficult end-of-life experiences identified in hospital settings, beyond those factors which we controlled for (22,24), or distress and anger amongst relatives whose loved ones died of terminal illness, without the expected levels of treatment or care (24). The slightly better levels of social support perceived amongst those who experienced care home deaths, compared with hospital and hospice deaths, might also help to explain this relationship (35–37).

### Circumstances of the death

There are a number of factors relating to the circumstances of the death that might be expected to have impacted upon grief levels. In previous quantitative and qualitative publications we identified experiences of sub-optimal end-of-life care, as healthcare systems and settings navigated the incredible strain and restrictions placed upon them (22,24). As in our baseline analysis (23), we again found that feeling supported by healthcare professionals immediately following the death had a significant and lasting positive effect on grief (PGD and IoV). Interestingly, we also found that this relationship with PGD scores was mediated by perceived social support at later time-points. This mediating effect might be explained by improved access to bereavement services as a result of supportive post-death care (and associated benefits relating to expressing feelings and receiving help with grieving, as captured in the ISS measure). It may also reflect the possibility that people with more negatives experiences (including problematic end-of-life care and related unanswered questions) may be more likely to feel poorly understood or unable to talk openly with others about how they are feeling, as also described in our qualitative findings (21,24). Alongside other pandemic (17,54,55) and non-pandemic studies (44,46) these findings demonstrate the importance of compassionate and effective communication around the time of death, and the likely significance of supportive post-death conversations and signposting for accessing further support and coming to terms with the circumstances of the death.

We found only small effects on PGD and IoV scores for factors relating to restricted contact at the end of life (e.g. visiting and saying goodbye), although being able to visit the patient at the end of life increased in importance once other factors were controlled in the PGD model. These relatively small effects are surprising given the devastating impacts of these experiences that were described in our qualitative data, including lasting feelings of guilt, anger, regret (24), and the effects of these circumstances identified in other studies (14,15,17,20). ‘Dealing with my feelings around how my loved one died’ was also the top-ranking need for support that we identified at baseline, with 60% of people experiencing high-level needs for help in this domain (21). Taken together these findings therefore suggest the significance of pandemic-related difficulties with end-of-life care and visiting relatives for grieving, in particular one’s ability to find meaning and come to terms with and accept the death (15,56).

### Disruption to grieving, coping and support processes

We found that loneliness and social isolation in early bereavement was strongly predictive of worse levels of PGD and IoV, as in our baseline analysis (23). Lower levels of social support at later time points were also strongly predictive of poorer PGD scores. These findings are consistent with pandemic and pre-pandemic evidence on the negative impacts of social isolation, loneliness and lack of social support on bereavement outcomes (6,29 35–37). Our qualitative findings provide detailed accounts of how lockdown restrictions and shielding not only limited the emotional support and comfort available to people, but also prevented the collective rituals and acts of remembrance needed to begin processing their grief (21,24). Whilst the negative impacts of poor social support on grief is not unique to the pandemic, this is a factor clearly exacerbated by the pandemic-context, as people suffered not only from physical separation but also emotional ‘distance’ and perceived lack of understanding and sensitivity to the realities of pandemic bereavement within social networks (15,17,21,23,24). Increased isolation and problematic social support during COVID-19 therefore seems likely to help explain elevated pandemic grief levels, whilst also reaffirming the important protective role of social support around the time of death and throughout bereavement generally (e.g.10, 35-37).

Although restricted funerals had a large effect on PGD (and medium effect on IoV) scores, this experience was not predictive once other factors were controlled for. This may be explained by the lessening effects of this experience over time and/or the greater relevance of other factors such as social support, along with the fact that almost all (93%) of respondents experienced these restrictions. Although most participants described the upsetting and distressing effects of restricted funerals in the qualitative data, a small minority described positive experiences and many described plans for future commemorative activities (24), which may have helped to mitigate the early effects of funeral restrictions. This uncertainty regarding the impact of funerals on bereavement outcomes is also reflected in the wider literature, which has been inconclusive (57) or found no effect (28), but also pointed to the important role of funeral providers and celebrants in providing alternative meaningful services in the contexts of restrictions.

### Demographic influences and participant characteristics

Several demographic or participant factors were found to have an effect on support and grief outcomes. Existence of medical conditions was associated with higher PGD (but not IoV) scores, and a small negative (but non-significant) effect on perceived social support. This points towards the detrimental impact of poorer health status on a person’s ability to cope and adjust, in particular at a time where clinically vulnerable populations were required to ‘shield’, and opportunities for usual social and recreational activities and access to services were heavily restricted (24,58). This is consistent with evidence from prior studies that existing mental-health conditions are associated with more complex grief (30,59) or poorer mental health outcomes (60). There is of course also the possibility that the bereavement itself may have led to new or worsened medical conditions amongst some participants, indicated in the increased numbers of people reporting new conditions across time. This would be consistent with studies reporting increased rates of morbidity and mortality amongst surviving spouses compared with general populations (e.g. the ‘widowhood’ effect, 61,62) and worse mental health amongst people bereaved during the pandemic compared with those not bereaved (60). Future analysis will investigate changes in health status and other associated factors over time (e.g. primary care and medication use) to explore these relationships further, along with other health economic outcomes, such as unemployment and time-off work.

As in previous research, and our baseline analysis (23), lower levels of education were associated with worse grief outcomes (PGD and IoV) (6,30,32–34). Although this factor became less important in the mixed-models, it underlines the importance of considering structural disadvantage and inequity in healthcare and bereavement support (27,63), particularly given the association that we previously identified between lower education-level and lower perceived healthcare professional support at the end of life (22). This effect might also relate to the unequal impacts of the pandemic on poorer communities across the UK, potentially affecting community-level mental health and resilience, and in turn a more limited capacity for healthy grieving and adaptation amongst people living in the worst affected localities. The overall negative mental health impact of living through the pandemic at general population-level has been documented (60,64,65), including worse outcomes associated with lower socio-economic status (6,65), economic stressors (66) and bereavement (60,66).

Despite the disproportionate impacts of the pandemic on minoritised ethnic communities in terms of death-rates, and disruption to grieving practices and community networks (27,67), this group of participants actually had better PGD (but not IoV) scores, although the difference was not statistically significant in the mixed-model. Further, the small size of this group (particularly at later time points) and the lessened effect of ethnicity once other factors were controlled, means that this finding should be treated cautiously. Of note though, is the observed potential mediating role of social support, and possibility that the better social support reported by our minority ethnic participants may have mitigated the effects of some of the general and culturally-specific challenges of pandemic bereavement faced by minoritised communities (e.g. see 21,27,64), although again these differences should be treated with caution.

### The bigger picture

This analysis has identified several pandemic-related factors which, in influencing grief outcomes, might at least partially explain the apparently higher levels of indicated PGD that we observed, compared with similar non-pandemic studies (7). When considered alongside our qualitative findings, however, what is also apparent is the intensity of feelings surrounding these and other factors; experiences which clearly had compounding and far-reaching effects on the lives of our participants, but which were not fully captured in our quantitative measures and analyses. Pandemic-related factors not fully measured or included in this analysis, but which we know to be highly consequential for grieving include: death-trauma (e.g. perceived suffering, poor treatment, shock), inability to collectively mourn or remember loved ones, the isolating and disenfranchising effects of being bereaved during a prolonged period of mass-bereavement (including lasting anger at political and societal responses to the pandemic and continuing fear of the virus), limited opportunities to engage in recreational and other coping activities, stressful death-administration and financial/work-based challenges, and reduced access to critical support-services (21, 24).

Within our qualitative findings, as in other studies (15), the significance of meaning-making in mediating the effects of many of these circumstantial factors is also evident. Examples of pandemic-related difficulties finding meaning included anger and unanswered questions surrounding and preventing acceptance of the death, descriptions of grief feeling ‘unreal’ without recourse to collective ritual, and lack of appropriate support and help with processing feelings (21,24). Pandemic-related disruption to meaning-making processes therefore may also explain the higher levels of pathological grief that we observed, as well as the small or insignificant effects of factors where only the occurrence of the ‘event’ rather than responses to it was captured (e.g. restricted funerals, being unable to say goodbye). This underlines the importance not only of considering how any infection-control restrictions that may be needed are managed and implemented (with meaningful alternatives available where possible), but equally that there is appropriate and effective communication with bereaved people and support surrounding any restrictions, coupled with opportunities to formally revisit and reflect upon what happened.

### Strengths and weaknesses

This longitudinal study benefits from a large initial sample-size, with quantitative and qualitative data collected across four time points up to approximately two years post-bereavement. Although participant numbers decreased over this time-period, we retained sufficient numbers to enable robust analysis, albeit with reducing proportions of younger and older participants, and people from minoritised ethnic backgrounds or with lowest qualifications levels. The sample was reasonably well represented across geographical areas, education and deprivation, but was self-selecting and biased towards female and white respondents, despite targeting men and people from minoritised ethnic communities in our recruitment approaches. By recruiting mostly online, we were also less likely to reach the very old or other digitally marginalised groups. Convenience sampling might have resulted in more people with negative experiences participating, as well as those accessing support. Despite these limitations, group sizes were sufficient to enable comparisons (although not to the level of specific ethnic groups) and, while not providing population-level prevalence data, the sample does enable comparisons to be made with data from similar pre-pandemic studies (e.g. 7), and the identification of potential risk factors which can inform future practice and policy.

### Implications for further research

Through subsequent qualitative interviews, we have explored in depth the experiences of people with characteristics less well represented in the survey, including men, people identifying with a sexual or ethnic minority background, with publication forthcoming. However, further research is required exploring the needs of bereaved people from minoritised ethnic backgrounds, same-sex couples, men, children and young people, and people with pre-existing mental health conditions (57), as we navigate the COVID-recovery phase and beyond. Given the importance of our qualitative data for establishing the ‘bigger picture’, the use of qualitative or mixed-methods approaches when investigating novel and unpredictable future mass-bereavement events is essential. The development or further refinement of tools for measuring identified event-specific risk factors e.g. the Inventory of Pandemic Grief Risk Factors (17), would also be helpful.

## CONCLUSIONS AND IMPLICATIONS FOR POLICY AND PRACTICE

We found relatively high-levels of indicated PGD at c. 8, 13 and 25 months post-bereavement when compared with similar non-pandemic studies of bereaved populations (e.g. 7). Several pandemic-related factors were identified which, in influencing grief outcomes, seem likely to at least partially explain this phenomenon. The strongest of these predictors were social isolation and problems accessing social support during bereavement, which whilst not unique to the pandemic was almost certainly exacerbated by it (21). Other likely explanatory factors included higher rates of unexpected deaths, and the disproportionately higher numbers of deaths occurring within socially deprived/less formally ‘educated’ communities during the pandemic (given the poorer grief outcomes of these groups). In their relationships with grief-levels, and the unique pandemic-context, poorer care- experiences at the end of life (including visiting restrictions) and the existence of other medical conditions, might also help to explain higher grief levels amongst people bereaved during the pandemic.

However, effect sizes for many of these factors were in absolute terms ‘small’, and our qualitative insights paint a much fuller and more intricate picture than we could capture in our quantitative measures and analyses. Taken together, our mixed-methods findings suggest that is likely the combined and compounding effects of the many different challenging experiences of people bereaved during the pandemic contributed to higher-levels of complex and prolonged grief.

Based on these findings we make the following recommendations to inform bereavement support and policy at the present time and in future pandemics, many of which resonate with the recent report by the UK Commission on Bereavement (67).

Implications for the current COVID-recovery phase and beyond:

1. In view of the higher proportions of people experiencing or at risk of PGD following the pandemic, bereavement support services require increased investment to ensure adequate levels of specialist provision which can effectively cater for those with more complex needs, as well as robust methods of identifying and reaching people most in need of more intensive support. Bereaved people more likely to require such support include those grieving children, partners and siblings and following unexpected deaths, as well as people who are isolated and have limited social support, health conditions and low levels of formal education.
2. Opportunities for informal emotional and social support should be strengthened through provision of peer-support groups, as well as compassionate community initiatives and educational programmes which seek to improve grief literacy and the support available to people within existing social and community networks. Communities worst affected by COVID-19 and structural inequalities should be prioritised for such initiatives.
3. Policies and training should be implemented to ensure compassionate and supportive communication and behaviours from healthcare professionals at the end of life, especially in acute and care-home settings. ‘Follow-up’ contact should be consistently delivered by care providers following the death and enable meaningful discussion and reflection on difficult and troubling experiences, with signposting to locally and nationally available bereavement support services.

To ensure preparedness for future pandemics and other mass-bereavement events, best practice- guidance and related policies should be developed for:

1. Health-care settings, with specific regard to managing and balancing infection-risk with the need to facilitate patient-family contact, including use of Personal Protective Equipment and remote communication-methods, and ensuring effective and compassionate communication with family members during times of crises.
2. Funeral-providers and crematoria, including identifying different options for meaningful and alternative funeral and mourning practices when restrictions are needed. The role of funeral directors in providing compassionate and supportive responses should be recognised, including their roles in sign-posting to further support-services (28, 54).
3. Managing social contact, recognising the need to restrict social interaction in times of high-infection rates, whilst making allowances for those living alone and with particular vulnerabilities, including the recently bereaved. Greater understanding of permissible levels of ‘safe’ contact relative to infection levels, and the best means of enabling this (e.g. outdoor socialisation) would also be helpful.
4. Rapid mobilisation of locally and nationally coordinated bereavement support provision, including existing providers and other community organisations. Any such responses should involve proactive sign-posting to and advertising of such support, mechanisms for identifying those requiring more intensive specialist support and crisis-specific training and practice- sharing to ensure that the support offered is crisis- as well as culturally-competent (21,68).

## Data Availability

Full study data (T1-T3) is available for download to users registered with the UK Data Service via https://reshare.ukdataservice.ac.uk/855751. T4 data (from the study extension) can be requested from lead/corresponding author.

## Acknowledgements

Our thanks to everyone who completed the survey for sharing their experiences, and to all the individuals and organisations that helped disseminate the survey. We would also like to thank the project assistants, collaborators and advisory group members who are not co-authors on this publication: Dr Anna Torrens-Burton, Dr. Emma Carduff, Dr. Daniella Holland-Hart, Prof. Bridget Johnston, Dr. Catriona Mayland, Dr. Donna Wakefield, Dr. Kirsten Smith, Dr. Audrey Roulston and Dr. Anne Finucane.

## Ethical standards

The authors assert that all procedures contributing to this work comply with the ethical standards of the relevant national and institutional committees on human experimentation and with the Helsinki Declaration of 1975, as revised in 2008.

## REFERENCES

1. WHO Coronavirus (COVID-19) Dashboard | WHO Coronavirus (COVID-19) Dashboard With Vaccination Data.

2. Menzies RE, Neimeyer RA, Menzies RG. Death anxiety, loss, and grief in the time of COVID- 19. Behaviour Change. 2020 Sep;37(3):111–5.

3. Palliative Care Australia. Palliative care and COVID19. Grief, bereavement and mental health. Palliative Care Australia. 2020

4. Eisma MC, Tamminga A. Grief before and during the COVID-19 pandemic: Multiple group comparisons. Journal of pain and symptom management. 2020 Dec 1;60(6):e1–4.

5. World Health Organization. International classification of diseases for mortality and morbidity statistics, 11th edn. 2018. https://icd.who.int/browse11/l-m/en [<u>Ref list</u>]

6. Shevlin M, Redican E, Hyland P, Murphy J, Karatzias T, McBride O, Bennett K, Butter S, Hartman TK, Vallières F, Bentall RP. Symptoms and levels of ICD-11 Prolonged Grief Disorder in a representative community sample of UK adults. Social Psychiatry and Psychiatric Epidemiology. 2023 Apr 11:1–3.

7. Aoun SM, Breen LJ, Howting DA, et al. Who needs bereave support? A population based survey of bereavement risk and support need. PLoS ONE 2015; 10(3): e0121101

8. Lundorff M, Holmgren H, Zachariae R, et al. Prevalence of prolonged grief disorder in adult bereavement: a system review and meta-analysis. J Affect Disord 2017; 212: 138–149. 6.

9. Prigerson HG, Horowitz MJ, Jacobs SC, et al. Prolonged grief disorder: psychometric validation of criteria proposed for DSM-V and ICD-11. PLoS Med 2009; 6(8): e1000121. 7.

10. Lobb EA, Kristjanson L, Aoun S, et al. Predictors of compli grief: a systematic review of empirical studies. Death Stud 2010; 34(8): 673–698.

11. Aoun SM, Breen LJ, O’Connor M, Rumbold B, Nordstrom C. A public health approach to bereavement support services in palliative care. Australian and New Zealand Journal of Public Health. 2012; 36: 14–16. pmid:22313700

12. Tang S, Xiang Z. Who suffered most after deaths due to COVID-19? Prevalence and correlates of prolonged grief disorder in COVID-19 related bereaved adults. Globalization and health. 2021 Dec;17(1):1-9.

13. Tang S, Yu Y, Chen Q, Fan M, Eisma MC. Correlates of mental health after COVID-19 bereavement in Mainland China. Journal of Pain and Symptom Management. 2021 Jun 1;61(6):e1–4.

14. Breen LJ, Lee SA, Neimeyer RA. Psychological risk factors of functional impairment after COVID-19 deaths. Journal of Pain and Symptom Management. 2021 Apr 1;61(4):e1–4.

15. Breen LJ, Mancini VO, Lee SA, Pappalardo EA, Neimeyer RA. Risk factors for dysfunctional grief and functional impairment for all causes of death during the COVID-19 pandemic: The mediating role of meaning. Death Studies. 2022 Jan 2;46(1):43–52.

16. Downar J, Parsons HA, Cohen L, Besserer E, Adeli S, Gratton V, Murphy R, Warmels G, Bruni A, Bhimji K, Dyason C. Bereavement outcomes in family members of those who died in acute care hospitals before and during the first wave of COVID-19: A cohort study. Palliative Medicine. 2022 Sep;36(8):1305–12.

17. Neimeyer RA, Lee SA. Circumstances of the death and associated risk factors for severity and impairment of COVID-19 grief. Death studies. 2022 Jan 2;46(1):34–42.

18. Gang, J., Falzarano, F., She, W.J., Winoker, H. and Prigerson, H.G., 2022. Are deaths from COVID-19 associated with higher rates of prolonged grief disorder (PGD) than deaths from other causes?. Death studies, 46(6), pp.1287–1296.

19. Eisma MC, Tamminga A, Smid GE, Boelen PA. Acute grief after deaths due to COVID-19, natural causes and unnatural causes: An empirical comparison. Journal of affective disorders. 2021 Jan 1;278:54–6.

20. Eisma MC, Tamminga A. COVID-19, natural, and unnatural bereavement: comprehensive comparisons of loss circumstances and grief severity. European Journal of Psychotraumatology. 2022 Jul 29;13(1):2062998.

21. Harrop E, Goss S, Farnell D, Longo M, Byrne A, Barawi K, Torrens-Burton A, Nelson A, Seddon K, Machin L, Sutton E. Support needs and barriers to accessing support: Baseline results of a mixed-methods national survey of people bereaved during the COVID-19 pandemic. Palliative medicine. 2021 Dec;35(10):1985–97.

22. Selman LE, Farnell DJ, Longo M, Goss S, Seddon K, Torrens-Burton A, Mayland CR, Wakefield D, Johnston B, Byrne A, Harrop E. Risk factors associated with poorer experiences of end-of-life care and challenges in early bereavement: Results of a national online survey of people bereaved during the COVID-19 pandemic. Palliative medicine. 2022 Apr;36(4):717–29.

23. Selman LE, Farnell DJ, Longo M, Goss S, Torrens-Burton A, Seddon K, Mayland CR, Machin L, Byrne A, Harrop EJ. Factors associated with higher levels of grief and support needs among people bereaved during the pandemic: Results from a national online survey. OMEGA-Journal of Death and Dying. 2022 Dec 21:00302228221144925.

24. Torrens-Burton A, Goss S, Sutton E, Barawi K, Longo M, Seddon K, Carduff E, Farnell DJ, Nelson A, Byrne A, Phillips R. ‘It was brutal. It still is’: a qualitative analysis of the challenges of bereavement during the COVID-19 pandemic reported in two national surveys. Palliative care and social practice. 2022 Apr;16:26323524221092456.

25. Harrop E, Selman LE. Bereavement during the Covid-19 pandemic in the UK: What do we know so far?. Bereavement Journal of Grief and Responses to Death. 2022 Jan 13;1.

26. Harrop E, Goss S, Longo M, Seddon K, Torrens-Burton A, Sutton E, Farnell DJ, Penny A, Nelson A, Byrne A, Selman LE. Parental perspectives on the grief and support needs of children and young people bereaved during the COVID-19 pandemic: qualitative findings from a national survey. BMC Palliative Care. 2022 Oct 10;21(1):177.

27. Selman LE, Sutton E, Medeiros Mirra R, Stone T, Gilbert E, Rolston Y, Murray K, Longo M, Seddon K, Penny A, Mayland CR. ‘Sadly I think we are sort of still quite white, middle-class really’–Inequities in access to bereavement support: Findings from a mixed methods study. Palliative Medicine. 2023 Apr;37(4):586–601.

28. Mitima-Verloop HB, Mooren T, Kritikou ME, Boelen PA. Restricted mourning: Impact of the COVID-19 pandemic on funeral services, grief rituals, and prolonged grief symptoms. Frontiers in psychiatry. 2022 May 27;13:1103.

29. Chen C, Tang S. Profiles of grief, post-traumatic stress, and post-traumatic growth among people bereaved due to COVID-19. European Journal of Psychotraumatology. 2021 Jan 1;12(1):1947563.

30. Newson RS, Boelen PA, Hek K, Hofman A, Tiemeier H. The prevalence and characteristics of complicated grief in older adults. Journal of affective disorders. 2011 Jul 1;132(1-2):231–8.

31. Ringdal GI, Jordhøy MS, Ringdal K, Kaasa S. Factors affecting grief reactions in close family members to individuals who have died of cancer. Journal of pain and symptom management. 2001 Dec 1;22(6):1016–26.

32. Mason TM, Tofthagen CS, Buck HG. Complicated grief: risk factors, protective factors, and interventions. Journal of social work in end-of-life & palliative care. 2020 Apr 2;16(2):151–74.

33. Milic J, Muka T, Ikram MA, Franco OH, Tiemeier H. Determinants and predictors of grief severity and persistence: the Rotterdam study. Journal of Aging and Health. 2017 Dec;29(8):1288–307.

34. Nielsen MK, Carlsen AH, Neergaard MA, Bidstrup PE, Guldin MB. Looking beyond the mean in grief trajectories: A prospective, population-based cohort study. Social Science & Medicine. 2019 Jul 1;232:460-

35. 35. Cacciatore J, Thieleman K, Fretts R, Jackson LB. What is good grief support? Exploring the actors and actions in social support after traumatic grief. PLoS One. 2021 May 27;16(5):e0252324.

36. 36. Stroebe, W., Zech, E., Stroebe, M. S., & Abakoumkin, G. (2005). Does social support help in bereavement? Journal of Social and Clinical Psychology, 24(7), 1030–1050.

37. 37. Smith KV, Wild J, Ehlers A. The masking of mourning: Social disconnection after bereavement and its role in psychological distress. Clinical Psychological Science. 2020 May;8(3):464-76.

38. Eysenbach G. Correction: improving the Quality of web surveys: the Checklist for Reporting results of internet E-Surveys (CHERRIES). Journal of medical Internet research. 2012 Jan 4;14(1):e2042.

39. 39. Boelen PA, Smid GE. The traumatic grief inventory self-report version (TGI-SR): Introduction and preliminary psychometric evaluation. Journal of Loss and Trauma. 2017 Apr 3;22(3):196–212.

40. Boelen PA, Djelantik AM, de Keijser J, Lenferink LI, Smid GE. Further validation of the Traumatic Grief Inventory-Self Report (TGI-SR): A measure of persistent complex bereavement disorder and prolonged grief disorder. Death studies. 2019 Jul 3;43(6):351–64.

41. Sim J, Machin L, Bartlam B. Identifying vulnerability in grief: psychometric properties of the Adult Attitude to Grief Scale. Quality of Life Research. 2014 May;23:1211–20.

42. 42. Machin, L. (2001). Exploring a framework for understanding the range of response to loss; a study of clients receiving bereavement counselling. . (Unpublished PhD thesis). Keele University, UK.

43. Hogan NS, Schmidt LA. Inventory of social support (ISS). InTechniques of grief therapy 2015 Sep 25 (pp. 99–102). Routledge.

44. Kentish-Barnes N, Chaize M, Seegers V, Legriel S, Cariou A, Jaber S, Lefrant JY, Floccard B, Renault A, Vinatier I, Mathonnet A. Complicated grief after death of a relative in the intensive care unit. European Respiratory Journal. 2015 May 1;45(5):1341–52.

45. Selman LE, Chao D, Sowden R, Marshall S, Chamberlain C, Koffman J. Bereavement support on the frontline of COVID-19: recommendations for hospital clinicians. Journal of pain and symptom management. 2020 Aug 1;60(2):e81–6.

46. Yamaguchi T, Maeda I, Hatano Y, Mori M, Shima Y, Tsuneto S, Kizawa Y, Morita T, Yamaguchi T, Aoyama M, Miyashita M. Effects of end-of-life discussions on the mental health of bereaved family members and quality of patient death and care. Journal of Pain and Symptom Management. 2017 Jul 1;54(1):17–26.

47. Haugen DF, Hufthammer KO, Gerlach C, Sigurdardottir K, Hansen MI, Ting G, Tripodoro VA, Goldraij G, Yanneo EG, Leppert W, Wolszczak K. Good quality care for cancer patients dying in hospitals, but information needs unmet: bereaved relatives’ survey within seven countries. The Oncologist. 2021 Jul;26(7):e1273–84.

48. Miyashita M, Morita T, Sato K, Hirai K, Shima Y, Uchitomi Y. Factors contributing to evaluation of a good death from the bereaved family member’s perspective. Psycho-Oncology: Journal of the Psychological, Social and Behavioral Dimensions of Cancer. 2008 Jun;17(6):612–20.

49. 49. R (version 4.1.1, R Core Team, 2021), R-Studio (www.r-studio.com).

50. Cohen, J. (1992). A power primer. Psychological Bulletin, 112, 155–159. doi:10.1037/0033-2909.112.1.155

51. Boelen PA, Smid GE, Mitima-Verloop HB, de Keijser J, Lenferink LI. Patterns, predictors, and prognostic validity of persistent complex bereavement disorder symptoms in recently bereaved adults: A latent class analysis. The Journal of Nervous and Mental Disease. 2019 Nov 1;207(11):913–20.

52. Dhada S, Stewart D, Cheema EA-O, Hadi MA-O, Paudyal VA-O. Cancer Services During the COVID-19 Pandemic: Systematic Review of Patient’s and Caregiver’s Experiences. 2021(1179–1322 (Print))

53. Lai AG, Pasea L, Banerjee A, Hall G, Denaxas S, Chang WH, et al. Estimated impact of the COVID-19 pandemic on cancer services and excess 1-year mortality in people with cancer and multimorbidity: near real-time data on cancer care, cancer deaths and a population-based cohort study. BMJ Open. 2020;10(11): e043828. doi: 10.1136/bmjopen-2020-043828

54. Mayland CR, Hughes R, Lane S, McGlinchey T, Donnellan W, Bennett K, Hanna J, Rapa E, Dalton L, Mason SR. Are public health measures and individualised care compatible in the face of a pandemic? A national observational study of bereaved relatives’ experiences during the COVID-19 pandemic. Palliative Medicine. 2021 Sep;35(8):1480–91.

55. Hanna JR, Rapa E, Dalton LJ, Hughes R, McGlinchey T, Bennett KM, Donnellan WJ, Mason SR, Mayland CR. A qualitative study of bereaved relatives’ end of life experiences during the COVID-19 pandemic. Palliative medicine. 2021 May;35(5):843–51.

56. Neimeyer RA. Meaning reconstruction & the experience of loss. American Psychological Association; 2001.

57. Burrell A, Selman LE. How do funeral practices impact bereaved relatives’ mental health, grief and bereavement? A mixed methods review with implications for COVID-19. OMEGA-Journal of Death and Dying. 2022 Jun;85(2):345–83.

58. Dos Santos CF, Picó-Pérez M, Morgado P. COVID-19 and mental health—what do we know so far?. Frontiers in Psychiatry. 2020 Oct 26;11:565698

59. Chiu, Y.W., Huang, C.T., Yin, S.M., Huang, Y.C., Chien, C.H. and Chuang, H.Y., 2010. Determinants of complicated grief in caregivers who cared for terminal cancer patients. Supportive Care in Cancer, 18, pp.1321–1327.

60. Joaquim RM, Pinto AL, Guatimosim RF, de Paula JJ, Costa DS, Diaz AP, da Silva AG, Pinheiro MI, Serpa AL, Miranda DM, Malloy-Diniz LF. Bereavement and psychological distress during COVID-19 pandemics: The impact of death experience on mental health. Current Research in Behavioral Sciences. 2021 Nov 1;2:100019.

61. A.R. Sullivan, A. Fenelon, Patterns of widowhood mortality, J. Gerontol.: Ser. Bibliogr. 69B (1) (2014) 53–62, https://doi.org/10.1093/geronb/gbt079.

62. Elwert F, Christakis NA. The effect of widowhood on mortality by the causes of death of both spouses. American journal of public health. 2008 Nov;98(11):2092–8.

63. Bindley K, Lewis J, Travaglia J, DiGiacomo M. Disadvantaged and disenfranchised in bereavement: A scoping review of social and structural inequity following expected death. Social Science & Medicine. 2019 Dec 1;242:112599.

64. Kira IA, Shuwiekh HA, Ashby JS, Elwakeel SA, Alhuwailah A, Sous MS, Baali SB, Azdaou C, Oliemat EM, Jamil HJ. The impact of COVID-19 traumatic stressors on mental health: Is COVID-19 a new trauma type. International Journal of Mental Health and Addiction. 2021 Jul 6:1–20.

65. Maffly-Kipp J, Eisenbeck N, Carreno DF, Hicks J. Mental health inequalities increase as a function of COVID-19 pandemic severity levels. Social Science & Medicine. 2021 Sep 1;285:114275.

66. Zacher M, Raker EJ, Meadows MC, Ramírez S, Woods T, Lowe SR. Mental health during the COVID-19 pandemic in a longitudinal study of Hurricane Katrina survivors. SSM-Mental Health. 2023 Dec 1;3:100198.Zacher et al.

67. UK Commission on Bereavement (2022) Bereavement is everybody’s business. Available at: https://bereavementcommission.org.uk/ukcb-findings/UKCB findings

68. Harrop E, Mann M, Semedo L, Chao D, Selman LE, Byrne A. What elements of a systems’ approach to bereavement are most effective in times of mass bereavement? A narrative systematic review with lessons for COVID-19. Palliative medicine. 2020 Oct;34(9):1165–81.

