## Supplementary file 2 for "Prolonged grief during and beyond the pandemic: Factors associated with levels of grief in a four time-point longitudinal survey of people bereaved in the first year of the COVID-19 pandemic"

### The grief experiences and support needs of people bereaved during Covid-19: Final Survey

Thank you once again for taking part in our study and agreeing to be sent this final questionnaire. We really appreciate the time you have taken over the last 18 months to help us with this study. Information from this final survey will be a great help in understanding longer-term experiences of grief and bereavement at this challenging time.

In this questionnaire we are interested to find out more about your grief experiences and wellbeing at the present time. We would also like to find out:

- What support you have been using recently.
- Any difficulties you may have experienced accessing support.
- What other type of support you feel you may still need.

More detailed information about the study is included in the document *Information for Participants – Final Survey* that is enclosed with your survey invitation. Please read this information to help you decide whether you would like to take part in this final survey and complete the consent section below if you would like to continue.

Information about bereavement support services and resources is also provided in the same document and at the end of this survey.

**Consent**

By participating in this survey, you agree that you have read and understood the information provided above and that you are aged 18 or over.

**I confirm that I have read and understood the information provided about the purpose of this study and how my data will be used, including the potential publication of my anonymised quotations to illustrate research findings. I agree to take part in the following survey knowing that all questions are optional and I can finish the survey at any.**

Please tick the box to agree with the above statement

**Thank you for your help.**

**Before we start, could you please tell us whether you have experienced any other bereavements of close friends or family members in the last year?**

| Yes |
| --- |
| No |

**If you have been bereaved again, we are deeply sorry to hear this and appreciate that you may not feel like completing this survey right now. If you would prefer for us to contact you in three months’ time instead please let us know below and use the enclosed prepaid envelope to return your survey to us [for online surveys: Please indicate below.].**

| I would prefer to complete this survey another time. |
| --- |
| I would like to continue with the survey. |

**Comments:**

**If you would like to continue, please carry on with the survey.**

**Part A**

This first section contains a series of questions which will help us to understand how your grief is affecting you at the moment and how you are coping with and adjusting to your bereavement. Some of these questions you may remember from answering previously, whilst others are new to this questionnaire. For all questions please remember that there are no right or wrong answers.

**A1. To help us better understand how you have been adjusting to the loss of your [insert], please indicate your response to the following attitudes:**

|  | **Strongly agree** | **Agree** | **Neither agree nor disagree** | **Disagree** | **Strongly disagree** |
| --- | --- | --- | --- | --- | --- |
| 1. I feel able to face the pain which comes with loss. | 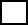 | 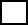 | 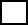 | 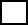 | 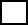 |
| 2. For me, it is difficult to switch off thoughts about the person I have lost. | 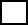 | 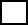 | 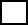 | 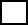 | 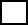 |
| 3. I feel very aware of my inner strength when faced with grief. | 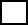 | 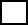 | 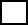 | 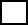 | 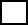 |
| 4. I believe that I must be brave in the face of loss. | 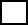 | 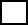 | 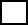 | 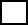 | 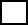 |
| 5. I feel that I will always carry the pain of grief with me. | 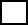 | 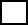 | 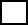 | 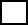 | 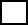 |
| 6. For me, it is important to keep my grief under control. | 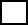 | 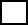 | 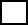 | 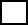 | 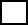 |
| 7. Life has less meaning for me after this loss. | 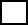 | 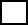 | 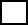 | 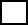 | 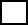 |
| 8. I think it’s best just to get on with life in spite of this loss. | 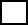 | 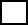 | 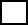 | 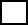 | 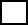 |
| 9. It may not always feel like it but I do believe that I will come through this experience of grief. | 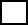 | 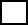 | 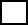 | 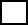 | 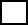 |

**Adult Attitude to Grief scale © Linda Machin 2000**

**A2. Several grief-reactions are listed below. Please indicate how often you have experienced each reaction in the past month, in response to the death of your [insert]:**

|  | **Never** | **Rarely** | **Sometimes** | **Frequently** | **Always** |
| --- | --- | --- | --- | --- | --- |
| 1. I had intrusive thoughts and images  related to the person who died. | 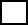 | 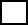 | 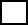 | 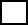 | 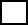 |
| 2. I experienced intense emotional pain,  sadness, or pangs of grief. | 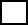 | 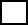 | 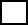 | 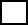 | 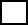 |
| 3. I found myself longing or yearning for the person who died. | 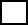 | 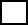 | 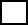 | 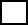 | 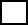 |
| 4. I experienced confusion about my role in life or a diminished sense of self. | 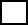 | 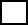 | 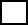 | 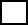 | 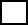 |
| 5. I had trouble accepting the loss. | 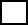 | 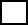 | 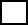 | 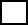 | 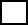 |
| 6. I avoided places, objects or thoughts  that reminded me that the person I lost has died. | 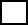 | 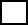 | 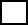 | 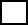 | 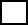 |
| 7. It was hard for me to trust others. | 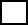 | 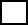 | 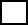 | 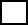 | 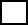 |
| 8. I felt bitterness or anger related to his/her death. | 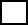 | 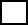 | 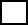 | 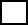 | 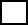 |
| 9. I felt that moving on (e.g. making new friends, pursuing new interests) was difficult for me. | 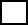 | 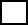 | 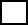 | 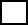 | 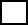 |
| 10. I felt emotionally numb. | 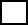 | 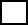 | 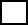 | 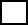 | 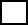 |
| 11. I felt that life is unfulfilling or meaningless without him/her. | 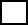 | 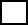 | 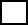 | 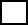 | 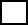 |
| 12. I felt stunned, shocked or dazed by his/her death. |  |  |  |  |  |
| 13. I noticed significant reduction in social, occupational, or other important areas of functioning (e.g., domestic responsibilities) as a result of his/her death. |  |  |  |  |  |
|  | **Never** | **Rarely** | **Sometimes** | **Frequently** | **Always** |
| 14. I had intrusive thoughts and images  associated with the circumstances of his/  her death. |  |  |  |  |  |
| 15. I experienced difficulty with positive reminiscing about the lost person. |  |  |  |  |  |
| 16. I had negative thoughts about myself in relation to the loss (e.g., thoughts about self-blame) |  |  |  |  |  |
| 17. I had a desire to die in order to be with the deceased. |  |  |  |  |  |
| 18. I felt alone or detached from other individuals. |  |  |  |  |  |

**Traumatic Grief Inventory-Self Report (Boelen et al. 2018)**

**A3. We would also like to get a sense of how well supported you feel at the moment. Using the scale shown below please select the statement that best describes the way you have been feeling during the past two weeks, including today.**

|  | **Does not describe me at all** | **Does not quite describe me** | **Describes me fairly well** | **Describes me well** | **Describes me very well** |
| --- | --- | --- | --- | --- | --- |
| People take the time to listen to how I feel |  |  |  |  |  |
| I can express my feelings about my grief openly and honestly |  |  |  |  |  |
| It helps me to talk with someone who is non-judgemental about how I grieve |  |  |  |  |  |
| There is at least one person I can talk to about my grief. |  |  |  |  |  |
| I can get help for my grieving when I need it. |  |  |  |  |  |

**Inventory for Social Support (Hogan and Schmidt 2002)**

**A4. Next we would like to ask you four questions about your feelings about aspects of your life. For each of these questions we would like you to give an answer on a scale of 0 to 10, where 0 is “not at all” and 10 is “completely”.**

|  | **Please rate each question on a scale from 0 (‘not at all’) to 10 (‘completely’)** | | | | | | | | | | |
| --- | --- | --- | --- | --- | --- | --- | --- | --- | --- | --- | --- |
|  | **0** | **1** | **2** | **3** | **4** | **5** | **6** | **7** | **8** | **9** | **10** |
| Overall, how satisfied are you with your life nowadays? |  |  |  |  |  |  |  |  |  |  |  |
| Overall, to what extent do you feel that the things you do in your life are worthwhile? |  |  |  |  |  |  |  |  |  |  |  |
| Overall, how happy did you feel yesterday? |  |  |  |  |  |  |  |  |  |  |  |
| On a scale where 0 is “not at all anxious” and 10 is “completely anxious”, overall, how anxious did you feel yesterday? |  |  |  |  |  |  |  |  |  |  |  |

**Source: Office National Statistics wellbeing measure**

**A5. If you would like to share a bit more on how you are currently coping with regards to the loss of your [insert], please use the box below.**

**Part B. Bereavement support**

**This section includes questions on your support needs and experiences of accessing support and help with your bereavement.**

**B1. Over the last TWO MONTHS have you needed support with the following?:**

|  | **High level of support needed** | **Fairly high level of support needed** | **Moderate level of support needed** | **Little support needed** | **No support needed** |
| --- | --- | --- | --- | --- | --- |
| Practical tasks relating to the death e.g. sorting out affairs, paperwork etc. |  |  |  |  |  |
| Getting relevant information and advice e.g. legal, financial, available support |  |  |  |  |  |
| Looking after myself/family e.g. getting food, medication, childcare etc |  |  |  |  |  |
| Dealing with my feelings about being without my loved one |  |  |  |  |  |
| Dealing with my feelings about the way my loved one died |  |  |  |  |  |
| Expressing my feelings and feeling understood by others |  |  |  |  |  |
| Feeling comforted and reassured |  |  |  |  |  |
| Loneliness and social isolation |  |  |  |  |  |
| Managing and maintaining my relationships with friends and family |  |  |  |  |  |
| Finding balance between grieving and other areas of life |  |  |  |  |  |
| Participating in work, leisure or other regular activities (e.g. shopping, housework) |  |  |  |  |  |
| Feelings of anxiety and depression |  |  |  |  |  |
| Regaining sense of purpose and meaning in life |  |  |  |  |  |

**Please provide further details on these or any other areas of support that you feel you have needed over the last six months:**

**B2. Over the last six months have you experienced any difficulties getting support for your grief and bereavement?**

|  | Yes | Somewhat | No | I’ve not tried to get their support |
| --- | --- | --- | --- | --- |
| From friends and family |  |  |  |  |
| From GP surgery |  |  |  |  |
| From bereavement services |  |  |  |  |

**B3. Do any of the responses below describe your experiences over the last six months? (Please select all that apply):**

| I have not wanted any support from bereavement services because my family and friends provide me with enough support |
| --- |
| I have not wanted any support from bereavement services because I am coping ok without this type of support |
| I have not wanted any support from bereavement services because I do not think it would help me |
| I do not know how to get support from bereavement services |
| I have felt uncomfortable asking for support from bereavement services |
| I have felt uncomfortable asking for help or support from friends or family |
| The support I wanted from bereavement services was not available to me |
| Friends or family have not been able to support me in the way I that wanted |

**B4. Please describe any difficulties that you have faced getting support from either friends/family or bereavement services in the last six months.**

**B5. Are there any children or young people under the age of 25 living with you who have also been affected by this bereavement?**

| Yes |
| --- |
| No |

**a. How old are they?**

**b. Please tell us about any support that you feel they need and/or any support they have been receiving during the last six months:**

**B6. Apart from any children or young people who might live with you, are there any other people close to you who have been particularly affected by this bereavement?**

**Section C. Types of support used**

**This section includes questions on the types of support you have been using over the last SIX months.**

**C1. What types of support or resources have you used over the last six months to help you cope with your bereavement**? (Please select all that apply)

| **Friend or family** |
| --- |
| **Written or audio resources** (e.g. self-help guides, books or websites, podcasts) |
| **GP or other member of staff at the GP surgery** |
| **Telephone helpline support** (e.g. bereavement helpline) |
| **Instant webchat support service** (e.g. exchanging written webchat messages with a trained or professional support provider in real time) |
| **Online bereavement community support via written comments** (e.g. Facebook group, online chat forums with other bereaved people) |
| **Community groups** (groups with a focus on socialising rather than bereavement support, e.g. faith groups, reading groups, gardening groups) |
| **Informal bereavement support group** (e.g. peer support group for bereaved people) |
| **Formal bereavement support group** (e.g. group discussions about bereavement guided by a trained or professional facilitator; or group counselling via video-call) |
| **One-to-one support** (e.g. individual counselling) |
| **Specialist mental health support** |

**If you selected “Community groups” above, how was this support provided?**

| In person |
| --- |
| Virtually via video/group calls |
| A mix of both |

**If you selected “Informal bereavement support group” above, how was this support provided?**

| In person |
| --- |
| Virtually via video/group calls |
| A mix of both |

**If you selected “Formal bereavement support group” above, how was this support provided?**

| In person |
| --- |
| Virtually via video/group calls |
| A mix of both |

**If you selected “One-to-one support” above, how was this support provided?**

| In person |
| --- |
| Virtually via video/group calls |
| A mix of both |

**If you selected “Specialist mental health support” above, how was this support provided?**

| In person |
| --- |
| Virtually via video/group calls |
| A mix of both |

**C2. Please tell us which of these type(s) of support you have found most helpful in the last six months and how it has helped you?**

**C3. Have you used any other types of support to those listed above?**

| Yes |
| --- |
| No |

**If yes please tell us what support this is and how it has helped you.**

**To help plan support services going forwards we would like to find out about your preferences for the different types of bereavement support that are available.**

**C4. Please tell us how much you would like or appreciate each of the following types of support if you were to experience another close bereavement in non-pandemic circumstances (i.e. when we can meet freely with others because infection control measures are no longer needed).**

1. **Support from Family and friends/Self-help resources**

|  | strongly like/quite like/neither like or dislike/slightly dislike/strongly dislike |
| --- | --- |
| **Friend or family** |  |
| **Written or audio resources** (e.g. self-help guides, books or websites, podcasts) |  |
| **Online bereavement community support via written comments** (e.g. Facebook group, online chat forums with other bereaved people) |  |

2. **Support from GP practice**

|  | strongly like/quite like/neither like or dislike/slightly dislike/strongly dislike |
| --- | --- |
| **Bereavement conversation with GP or other member of practice staff** (including signposting and/or timely referrals to support) |  |

3. **Instant access bereavement support**

|  | strongly like/quite like/neither like or dislike/slightly dislike/strongly dislike |
| --- | --- |
| **Telephone helpline support** (e.g. bereavement helpline) |  |
| **Instant webchat support service** (e.g. exchanging written webchat messages with a trained or professional support provider in real time) |  |

4. **Group-based support (both in person or virtual support via video calls)**

|  | strongly like/quite like/neither like or dislike/slightly dislike/strongly dislike |
| --- | --- |
| **Community groups** (groups with a focus on socialising rather than bereavement support, e.g. faith groups or reading groups, gardening groups) **– IN PERSON SUPPORT** |  |
| **Community groups** (groups with a focus on socialising rather than bereavement support, e.g. faith groups, reading groups) **– VIRTUAL SUPPORT VIA VIDEO CALLS** |  |
| **Informal bereavement support group** (e.g. peer support group for bereaved people) **-** **IN PERSON SUPPORT** |  |
| **Informal bereavement support group** (e.g. peer support group for bereaved people) **– VIRTUAL SUPPORT VIA VIDEO CALLS** |  |
| **Formal bereavement support group** (e.g. group discussions about bereavement guided by a trained or professional facilitator; or group counselling) **– IN PERSON SUPPORT** |  |
| **Formal bereavement support group** (e.g. group discussions about bereavement guided by a trained or professional facilitator; or group counselling) **– VIRTUAL SUPPORT VIA VIDEO CALLS** |  |

5. **One-to-one support**

|  | strongly like/quite like/neither like or dislike/slightly dislike/strongly dislike |
| --- | --- |
| **One-to-one support** (e.g. individual counselling) – **PROVIDED IN PERSON** |  |
| **One-to-one support** (e.g. individual counselling) – **PROVIDED VIA TELEPHONE OR VIDEO CALL** |  |
| **Specialist mental health support** – **PROVIDED IN PERSON** |  |
| **Specialist mental health support** – **PROVIDED VIA TELEPHONE OR VIDEO CALL** |  |

**C5. Please use the box below to tell us more about your preferences for these different types of support.**

**C6. Thinking back on the time since the loss of xxx , what aspect(s) of your bereavement or grief have you found the most challenging?**

**C7. What helped or would have helped you the most, to cope and adjust over this period of time?**

**Section D.**

**In this section we would like to ask you a few additional questions about other aspects of your life that may affect or have been affected by your bereavement.**

**D1. Have you been diagnosed with any illness or medical condition in the past year?**

| Yes |
| --- |
| No |

If yes, please provide details:

……………………………………………………………………………………………………………………………………………….

**D2. Roughly how many appointments with your GP have you had over the last two months?**

……………………………………………………………………………………………………………………………………………….

**D3. Roughly how many times have you bought over-the-counter medicine (e.g. paracetamol, sleep aids or stress relief remedies) over the last two months?**

| Never |
| --- |
| One to three times |
| 4 times or more |

**D4. Have you experienced any difficulties with sleeping over the last two months?**

| Yes |
| --- |
| No  *If no, please continue to question E5.* |

**If yes, how often do you have difficulties with sleeping (e.g. trouble falling asleep, waking up early or in the middle of the night, bad dreams, poor overall sleep quality)?**

| Less than once a week |
| --- |
| Once or twice a week |
| Three or more times a week |

**To enable us to better understand how bereavement impacts upon the employment and** **working life of bereaved people, please answer the following questions.**

**D5. Has your employment status changed in the last 12 months?**

| **Yes** |
| --- |
| **No** |
| **Not relevant for me**  *(e.g. retired, full-time student, permanently sick/disabled, long-term unemployed, looking after the home, caring for a loved one)*  **If not relevant for you, please continue to the end of the survey on page 21.** |

If yes, please provide details (including if you have been furloughed):

……………………………………………………………………………………………………………………………………………….

**D6. Have you had any time off work during the last SIX MONTHS due to bereavement, illness or stress?**

| **Yes** |
| --- |
| **No** |
| **Not relevant for me**  *(e.g. retired, full-time student, permanently sick/disabled, long-term unemployed, looking after the home, caring for a loved one)* |

**If yes, how much time have you had off work due to bereavement, illness or stress (in days/weeks or months?)**

| Amount of time off work due to bereavement, illness or stress: |
| --- |

**D7. If you would like to share further information about your time off work in the last six months, please use the box below:**

**Many thanks for completing this final survey and for all of the time that you have given to this research.** Your responses will help to enable others who are bereaved to access the support they need. We appreciate that this may have been difficult and painful for you and we are very grateful for your contribution.

Thank you again for your help. We are extremely grateful for your contribution.

If you would like to talk to somebody about your bereavement, you can access support from these services:

- Marie Curie Bereavement Support: 0800 090 2309 <https://www.mariecurie.org.uk/help/support/bereaved-family-friends/dealing-grief/bereavement-or-grief-counselling>
- Cruse Bereavement Care: 0808 808 1677

<https://www.cruse.org.uk/>

- NHS Bereavement Helpline: 0800 2600 400 <https://www.nhs.uk/conditions/stress-anxiety-depression/coping-with-bereavement/>
- The Good Grief Trust: <https://www.thegoodgrieftrust.org/>
- At a Loss: [www.ataloss.org](http://www.ataloss.org)

***Thank you again for your help.***
